# UB-612 Multitope Vaccine Targeting SARS-CoV-2 Spike and Non-Spike Proteins Provides Broad and Durable Immune Responses

**DOI:** 10.1101/2022.08.26.22279232

**Authors:** Chang Yi Wang, Wen-Jiun Peng, Be-Sheng Kuo, Hope Liu, Yu-Hsin Ho, Min-Sheng Wang, Ya-Ting Yang, Po-Yen Chang, Yea-Huei Shen, Kao-Pin Hwang

## Abstract

The SARS-CoV-2 non-Spike (S) structural protein targets of nucleocapsid (N), membrane (M) and envelope (E), critical in the host cell interferon response and memory T-cell immunity, have been grossly overlooked since the inception of COVID vaccine development. To pursue a universal (pan-sarbecovirus) vaccine against ever-emergent future mutants, we explored booster immunogenicity of UB-612, a multitope-vaccine that contains S1-RBD-sFc protein and sequence-conserved rationally designed promiscuous Th and CTL epitope peptides on the Sarbecovirus N, M and S2 proteins. To a subpopulation of infection-free participants (aged 18-85 years) involved in a two-dose Phase-2 trial, a UB-612 booster (third dose) was administered 6-8 months after the second dose. The immunogenicity was evaluated at 14 days post-booster with overall safety monitored until the end of study. The booster induced high viral-neutralizing antibodies against live Wuhan WT (VNT_50_, 1,711) and Delta (VNT_50_, 1,282); and against pseudovirus WT (pVNT_50,_ 11,167) vs. Omicron BA.1/BA.2/BA.5 variants (pVNT_50_, 2,314/1,890/854), respectively. The lower primary neutralizing antibodies in the elderly were uplifted upon boosting to approximately the same high level in young adults. UB-612 also induced potent, durable Th1-oriented (IFN-γ^+^-) responses (peak/pre-boost/post-boost SFU/10^6^ PBMCs, 374/261/444) along with robust presence of cytotoxic CD8^+^ T cells (peak/pre-boost/post-boost CD107a^+^-Granzyme B^+^, 3.6%/1.8%/1.8%). Booster vaccination is safe and well tolerated without SAEs. By recognition against epitopes on Spike (S1-RBD and S2) and non-Spike (N and M) structure proteins, UB-612 provides potent, broad and long-lasting B-cell and T-cell memory immunity and offers a potential as a universal vaccine to fend off Omicrons and new VoCs.

**SIGNIFICANCE STATEMENT:** The Omicron has swept the globe with a rapid succession of dominating sublineages from BA.1, BA.2, to the current BA.5 with increasing infectivity and antibody evasion. Concerningly, the non-Spike structure proteins that promote T-cell immunity are grossly overlooked in vaccine development. Looking beyond short-interval booster jabs and omicron-updated vaccines, a pragmatic approach to curbing ever-emergent new mutants would be “universal (pan-Sarbecovirus) vaccines” targeting conserved nonmutable epitopes on coronavirus. UB-612, a multitope-vaccine armed with Spike (S1-RBD and S2) and non-Spike targets (Nucleocapsid N and Membrane M), allows booster vaccination to elicit potent, broadly-recognizing, durable B- and T-cell memory immunity. Sequence-conserved epitope peptides were rationally-designed from S2, N and M proteins to synergistically enhance memory helper and cytotoxic T-cell immunity and B-cell immunity.

## INTRODUCTION

The SARS-CoV-2 Omicron lineage has swept the globe with a rapid succession of dominating subvariants from BA.1, BA.2 and to the current BA.5 that takes up more than 90% infection cases with overriding edges in transmissibility and neutralizing antibody escape (1-7). Relative to Delta variant, Omicron BA.1 does not require fusion co-receptor TMPRSS2 for cell entry; instead, cells engulf it and land intracellularly through endosomes (8, 9). It infects upper bronchial cells and proliferates much more efficiently; it does not foster cell syncytia nor erode lung-alveolar tissues (8-14), thus causing lesser disease severity.

Omicron BA.1 is heavily mutated (**Table 1A**), including 35 changes in S protein (8, 15-17). Compared to 2 mutations associated with Delta at S-1 receptor binding domain (S1-RBD, residues 319-541), BA.1 and BA.2 share 12 mutations, with BA.1 and BA.2 each having additional 3 and 4 unique ones, respectively, that confers BA.2 a higher immune escape.

**Table 1.**
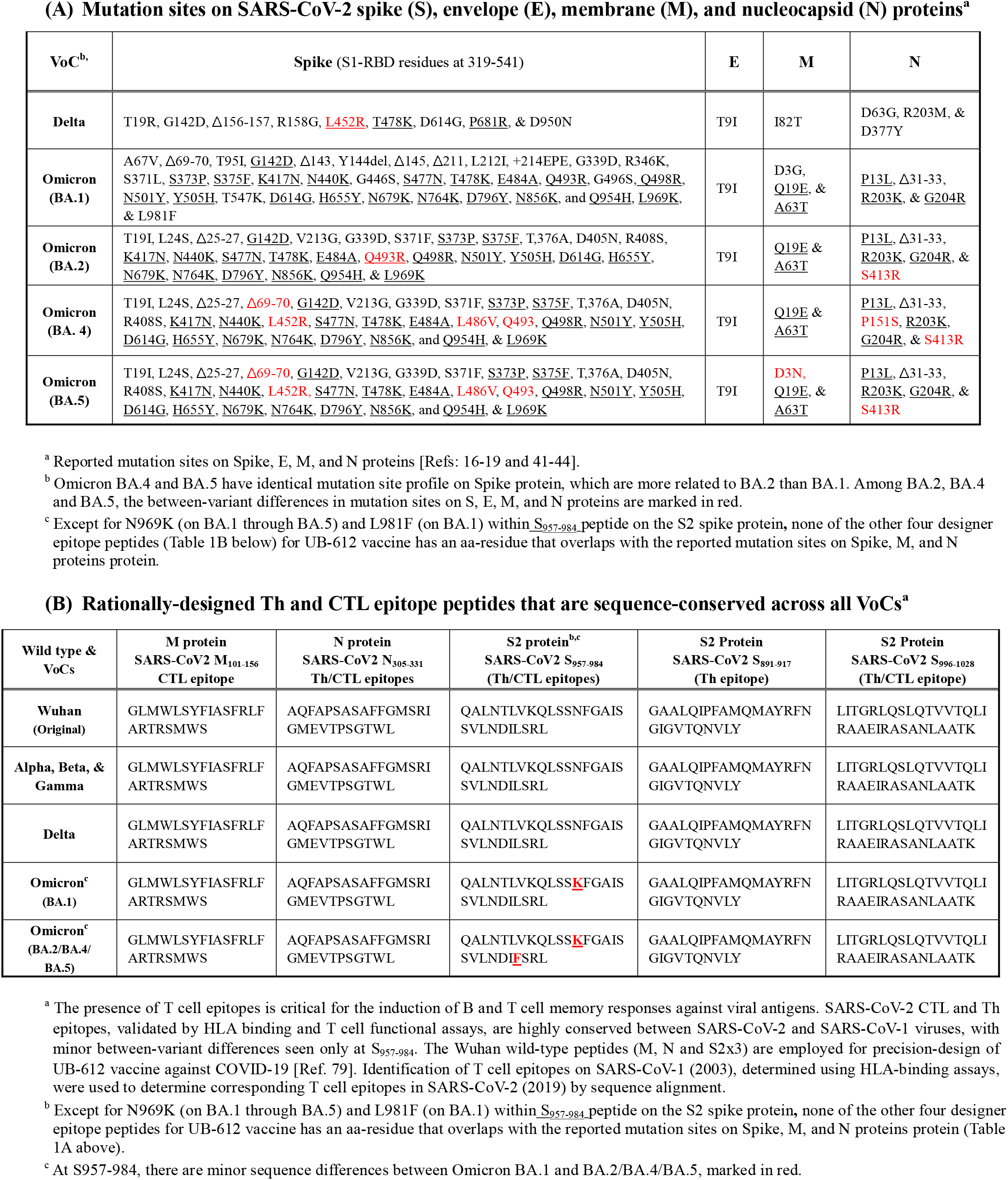
The mutation sites on SARS-CoV-2 Spike (S), Envelope (E), membrane (M) and nucleocapsid (N) proteins on Delta and Omicrons and the rationally-designed sequence-conserved Th/CTL epitope peptides on M, N, and S2 proteins across all Variants of Concern (VoC)

BA.4 and BA.5 have identical spike protein. They differ from BA.2 by having additional mutations at 69-70del, L452R, F486V and wild type amino acid at position Q493 (18) (**Table 1A**), contributing to their higher degree of immune escape than BA.2.

BA.2 exhibits a 1.3- to 1.5-fold higher transmissibility and a 1.3-fold immune escape than BA.1 (16, 19), consistent with the finding that BA.1-immune sera neutralizes BA.2 with lower titers by a factor of 1.3 to 1.4 (20) and that BA.2 reinfection can occur after BA.1 (21). BA.4/BA.5 are more transmissible and resistant to BA.1/BA.2-immunity and monoclonal antibodies (1, 2). While BA.2 vs. BA.1 (19) and BA.4/BA.5 vs. BA.2 (22) display greater cell-to-cell fusion, they do not make infected people sicker nor change the fundamental pandemic dynamics.

Of double-vaccinated adults, the booster (third dose)-induced neutralization titers against BA.4/BA.5 are notably lower than those against BA.1/BA.2 (3-5); and, of BA.1-infected adults, the immune sera can potently neutralized BA.1/BA.2, but weaker against BA.4/BA.5 (6, 22, 23). These suggest that booster vaccination or BA.1/BA.2 infection may not achieve sufficient immunity to protection against BA.4/BA.5. And, regardless of vaccination status or hybrid immunity, each reinfection would add risks of mortality, hospitalization and other health hazards including burden of long-haul COVID (24).

In the real-world, while a booster 3^rd^-dose of mRNA vaccines could compensate Omicron (BA.1)-induced decrease of serum neutralizing antibodies (20- to 30-fold reduction) and reduce rates of hospitalization and severe disease (80-90% protection) (25-30), they offer less effective protection against mild and asymptomatic infections (40-50% protection) (30). Breakthrough infections (mostly asymptomatic and mild) tagged with high viral loads are common even after the fourth jab (2^nd^ booster to adults aged 18 and older) (31, 32), which offers only marginal benefits against severe disease and the effectiveness against infection (asymptomatic & mild cases) was as low as 11-30%. These suggest the magnitude and the durability of vaccine-induced immunity by the currently licensed COVID vaccines remain a key issue to be addressed.

While development of composition-updated (variant-specific) vaccines has been strongly advocated (33, 34), a better yet strategy of “universal coronavirus vaccines” would be in a more urgent need (35) for robust, broad, and durable immunity. The currently authorized Spike-only vaccines do not incorporate SARS-CoV-2’s non-spike structure proteins of envelope (E), membrane (M) and nucleocapsid (N), the regions critically involved in the host cell interferon response and T-cell memory (36-38). Oversight of non-Spike proteins as targets could lead to an intrinsic shortfall for promotion of a fuller T cell immunity. Viral mutations are also known to occur in E, M and N **(Table 1A)** (15-18, 39-42), the structure proteins that are beyond recognition by current vaccines.

In the present Phase-2 extension study, we affirm that UB-612 booster (third dose) can induce potent, broadly-recognizing, durable antibodies and T-cell immunity that offers a potential as a universal vaccine to fend off Omicrons and new mutants. UB-612 is a recently EUA approved multitope-vaccine which contains S1-RBD-sFc fusion protein enriched with five rationally-designed promiscuous peptides representing sequence-conserved Th and CTL epitopes on the Sarbecovirus N, M and S2 proteins across all VoCs (**Table 1B**) and a sixth idealized universal Th peptide which serves as a catalyst in T cell activation.

## METHODS

### Design of Phase-2 Extension Booster Trial and Oversight

#### Booster 3rd-dose following the Phase-2 trial primary 2-dose series

We conducted a booster vaccination study (n = 1,478) which was an extension arm of the Phase-2, placebo-controlled, randomized, observer-blind, multi-center primary 2-dose study (**Supplemental Figure 1A**) [ClinicalTrials.gov: NCT04773067] in Taiwan with 3,844 healthy male or female adults aged >18 to 85 years (**Supplemental Figure 1B**) who received two intramuscular doses (28 days apart) of 100 μg UB-612 or saline placebo. The objectives of the third-dose extension study were to determine the booster-induced safety and immunogenicity after unblinding, 6 to 8 months after the second dose.

The Principal Investigators at the study sites agreed to conduct the study according to the specifics of the study protocol and the principles of Good Clinical Practice (GCP); and all the investigators assured accuracy and completeness of the data and analyses presented. The protocol was approved by the ethics committee at the sites and all participants provided written informed consent. Full details of the booster trial design, inclusion and exclusion criteria, conduct, oversight, and statistical analysis plan are available in the study protocol in ***Supplemental Information***.

### Trial Procedures of Safety and Immunogenicity

#### Reactogenicity in the primary and booster series

The primary safety endpoints of the Phase-2 primary series (Days 1-365) and extension booster trial (recorded until 14 days post-booster and followed up study end) were to evaluate the safety and tolerability. Vital signs were assessed before and after each injection. After each injection, participants had to record solicited local and systemic AEs in their self-evaluation e-diary for up to seven days while skin allergic reactions were recorded in their e-diary for up to fourteen days. Safety endpoints include unsolicited AEs reported for Days 1 to 57 in primary series and Day 1 to 14 in the booster phase. The overall safety was followed until the end of this study. Complete details for solicited reactions are provided in the study protocols.

#### Scope of immunogenicity investigation

The primary immunogenicity endpoints were the geometric mean titers (GMT) of neutralizing antibodies against SARS-CoV-2 wild-type (WT, Wuhan strain), Delta, Omicron BA.1, BA.2 and BA.5 variants were explored. For WT and Delta strains, viral-neutralizing antibody titers that neutralize 50% (VNT50) of live SARS-CoV-2 WT and Delta variant were measured by a cytopathic effect (CPE)-based assay using Vero-E6 (ATCC® CRL-1586) cells challenged with SARS-CoV-2-Taiwan-CDC#4 (Wuhan strain) and SARS-CoV-2-Taiwan-CDC#1144 (B.1.617.2; Delta variant). The replicating virus neutralization test conducted at Academia Sinica was fully validated using internal reference controls and results expressed as VNT_50_. For WT, Omicron BA.1, BA.2, and BA.4/BA.5 strains, 50% pseudovirus neutralization titers (pVNT_50_) were measured by neutralization assay using HEK-293T-ACE2 cells challenged with SARS-CoV-2 pseudovirus variants expressing the spike protein of WT, BA.1, BA.2, or BA.4/BA.5 variants.

The secondary immunogenicity endpoints include anti-S1-RBD IgG antibody, inhibitory titers against ACE2:RBDWT interaction, and T-cell responses assayed by ELISpot and Intracellular Staining. The RBD IgG ELISA was fully validated using internal reference controls and results expressed in end-point titers. A panel of 20 human convalescent serum samples from COVID-19 Taiwan hospitalized patients aged 20 to 55 years were also tested for comparison with those in the vaccinees. Human peripheral blood mononuclear cells (PBMCs) were used for monitoring T cell responses (ELISpot and ICS). The constructs of the UB-612 vaccine product, all bioassay methods and statistics are detailed in the ***Supplemental Methods***.

## RESULTS

### Booster trial population

After unblinding of Phase-2 trial, 1,478 of the 3,844 healthy study participants who completed the 2-dose primary vaccine series (28 days apart) of 100-μg UB-612 (**Supplemental Figure 1A**) were enrolled to receive an additional 100-μg booster 3rd-dose at 6 to 8 months after the second shot. The booster vaccinees were followed for 14 days to evaluate safety and immunogenicity. The vast majority of participants were of Taiwanese origin, with two groups aged 18-65 years (76%) and 65-85 years (24%) (**Supplemental Figure 1B**).

### Reactogenicity and safety

No vaccine-related serious adverse events (SAEs) were recorded; the most common solicited AEs were injection site pain and fatigue, mostly mild and transient (**Supplemental Figure 2**). The incidence of solicited local AEs slightly increased at post-booster (**Supplemental Figure 2A**), mostly pain at the injection site (mild, 54%; moderate, 7%). The incidence of skin allergic reaction at post-booster was similar to post-dose 2 (**Supplemental Figure 2B**). Fatigue/tiredness, muscle pain, and headaches that belonged to solicited systemic AEs were mostly mild (**Supplemental Figure 2C**). Overall, no safety concerns were identified with UB-612 booster across age groups.

### Durable Th1 cell responses by ELISpot

Vaccinees’ peripheral blood mononuclear cells (PBMCs) were collected for evaluation of Interferon-γ^+^ (IFN-γ^+^)-ELISpot. On Day 57 (28 days post-2^nd^ dose), IFN-γ SFU (Spot Forming Unit)/10^6^ cells under stimulation with RBD+Th/CTL peptide pool (**Figure 1A**) increased from the baseline 1.0 to a high peak at 374 SFU/10^6^, which maintained robust at 261 (70%) at pre-boosting (6-8 months post-2^nd^ dose) and rose to 444 SFU 14 days post-booster (**Figure 1D**).

**Figure 1.**
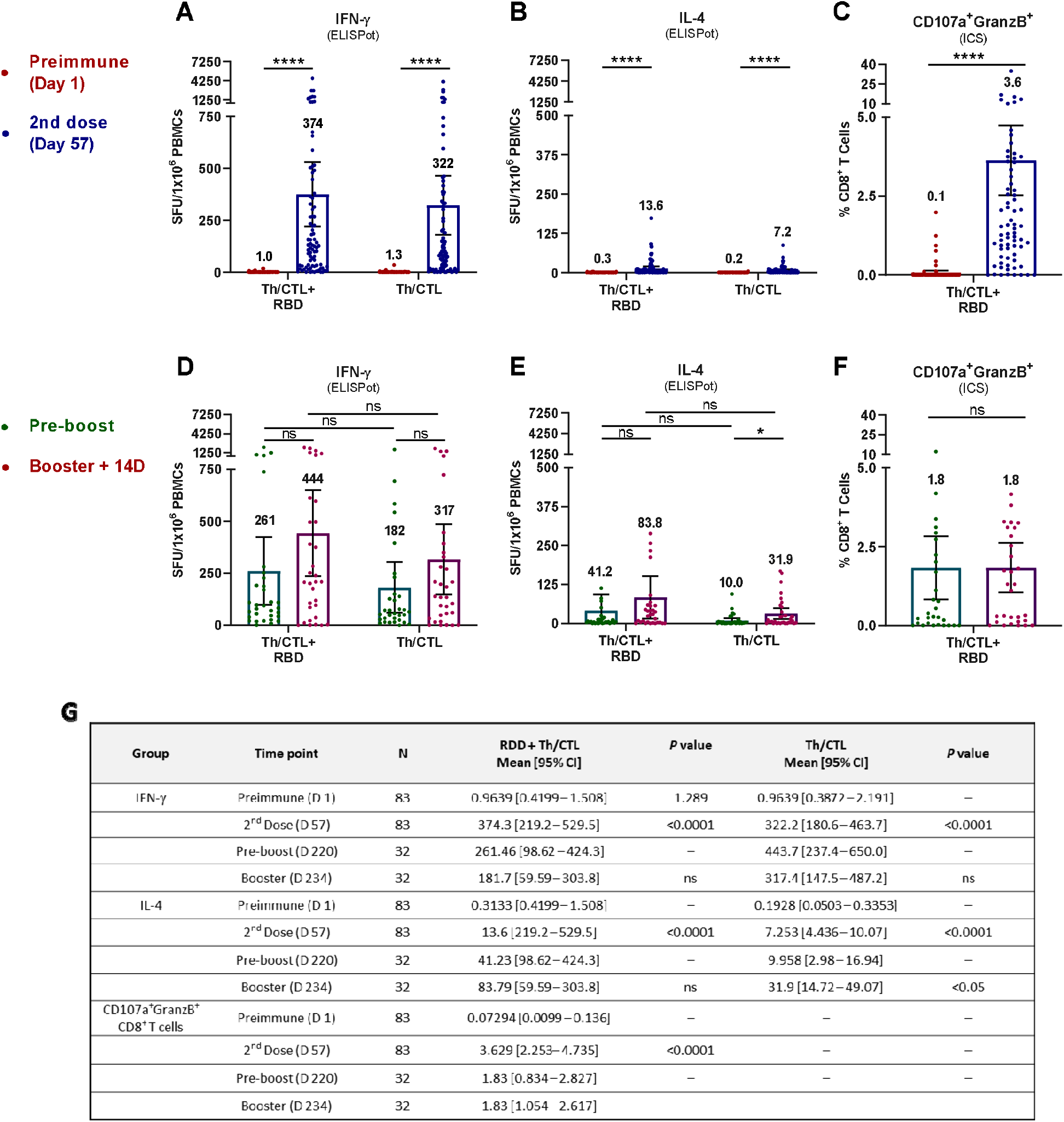
UB-612 induced T cell responses measured by ELISpot and ICS analyses. T-cell responses to stimulation by epitope peptides (RBD+Th/CTL or Th/CTL alone) were analysed with PBMCs collected from 83 vaccinees from Immunogenicity group (n = 83) on Days 57 (28 days after 2^nd^ booster); and from 32 vaccinees from the Immunogenicity (n = 18) or Safety groups (n = 14) who joined the Phase-2 extension booster study to evaluate the T-cell responses in PBMCs on Days 197 to 242 (pre-boosting days) and Days 211 to 256 (14 days post-booster third dose). T-cell responses were measured by **(A, D)** IFN-γ and **(B, E)** IL-4 ELISpot at 10-μg/mL per stimulator. Spot-forming units (SFU) per 1×10^6^ PBMCs producing IFN-γ and IL-4 after stimulation with the RBD+Th/CTL peptide pool or the Th/CTL peptide pool are expressed. The PBMC samples stimulated with Th/CTL+RBD were also evaluated for T cell responses by Intracellular Staining (ICS) (**C, F**), by which the frequency of CD8^+^ T cells producing the measured cytokines (CD107a and Granzyme B) in response to the stimulation by RBD+Th/CTL peptide pool is shown. (**G**) Summary of mean and 95% CI are presented for plots as shown in Figures **(A)** to **(F)**. Horizontal bars indicate mean with 95% CI.

Similar IFN-γ profiles were observed for those stimulated with Th/CTL peptide pool alone (**Figure 1A**), which increased from the baseline 1.3 to a high peak at 322 SFU/10^6^ cells on Day 57, maintained at 182 SFU/10^6^ cells (∼57%) at pre-boosting and remained strong at 317 SFU/10^6^ cells 14 days post-booster (**Figure 1D**). T cell responses persisted robustly (60-70% of the high peak at Day 57) long over 6-8 months.

These results indicate that UB-612 can induce a strong and durable IFN-γ^+^ T cell immunity in the primary series, prompt a high level of memory recall upon boosting, and the presence of Th/CTL peptides is essential and principally responsible for the bulk of the T cell responses, while S1-RBD domain plays a minor role. Together with the insignificant low levels of the IL-4^+^ ELISpot responses (**Figure 1, B and E**), UB-612 vaccination as both primary series and homologous-boosting could induce pronounced Th1-predominant immunity.

### High CTL CD8^+^ T cell activity by Intracellular Cytokine Staining (ICS)

Along with high levels of ELISpot-based T cell responses, the vaccine recipients also showed cytotoxic T-cell responses, including CD8^+^ T cells expressing cytotoxic markers CD107a and Granzyme B.as observed in the primary series, accounting for a remarkable 3.6% of circulating CD8^+^ T cells after re-stimulation with S1-RBD+Th/CTL peptide pool (**Figure 1C**), which persisted at a substantial 1.8% upon booster vaccination (**Figure 1F**). Apparently, CD8^+^ T cell responses persisted robustly (50% of the high peak at Day 57) over 6-8 months as well. This suggests a potential of robust cytotoxic CD8^+^ T responses in favor of viral clearance once infection occurs.

### Overview of B cell immunogenicity on antigenic and functional levels

Of the 871 Phase-2 study participants designated for Immunogenicity investigation, 302 participants had their serum samples collected at pre-boosting and 14 days post-booster for antigenic assay by anti-S1-RBD IgG ELISA, and functional assays by ACE2:RBD_**WT**_ binding inhibition ELISA and by neutralization against live SARS-CoV-2 wild-type Wuhan strain (WT) by cell-based CPE method (**Supplemental Figure 3**). The results showed pronounced booster-induced increase of antibody titers that bound to RBD and inhibit/neutralize ACE2 interaction by respective 16- to 45-folds. These indicate that UB-612 booster vaccination could profoundly enhance both antigenic and functional activities.

### Neutralizing antibodies against WT, Delta, Omicron BA.1 and BA.2

Functional blockade was further investigated comparatively on the occasion when Omicrons BA.1 and BA.2 dominate the pandemic scene. First, with limited available, affordable sources of viral variants, we investigated immune sera from 41 study participants across all age groups (18-65 years, n = 26; 65-85 years, n = 15). Neutralization measured using live virus, UB-612 booster elicited a neutralizing titer (VNT_50_) against WT at 1,711 versus Delta variant at 1,282 (**Figure 2A**), representing a 1.3-fold reduction (GMFR, Geomean Fold Reduction). There was no significant age-dependent neutralization effect between young adults (18-65 yrs.) and the elderly (65-85 yrs.) with respect to either anti-WT or anti-Delta VNT_50_ levels (**Figure 2B**), with a modest 1.2- to 1.7-fold GMFR of anti-Delta relative to anti-WT level.

**Figure 2.**
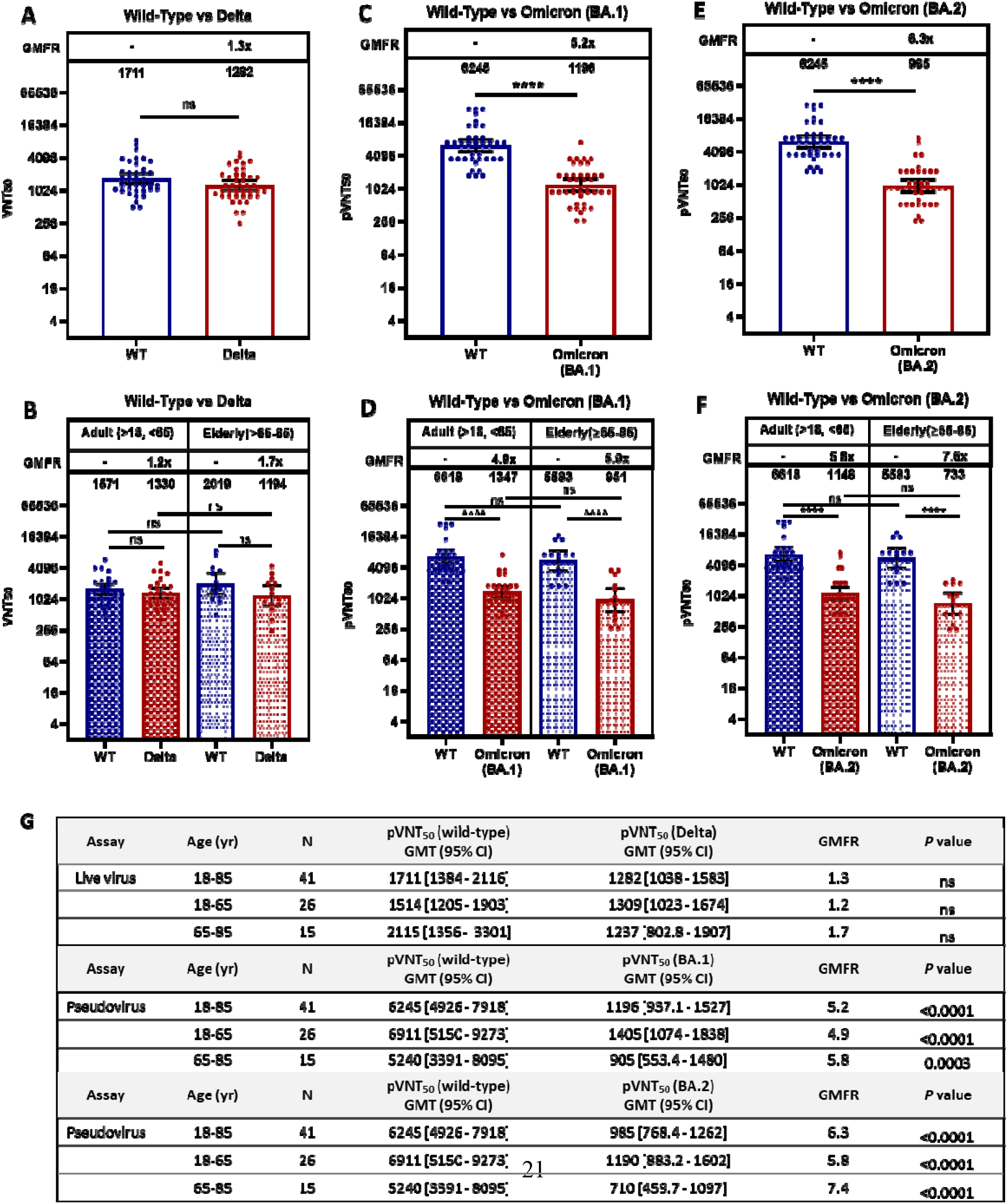
Viral-neutralization effects of UB-612 booster vaccination against wild type, Delta, Omicron BA.1 and BA.2 variants. Viral-neutralizing titers against SARS-CoV-2 wild-type, Delta, Omicron BA.1, and BA.2variants were investigated during the infection pandemic that BA.1/BA dominated. Serum samples from 41 participants (n = 27 for 18-65 years; n = 14 for 65-85 years) collected at 14 days post-booster were subjected to a live virus or pseudovirus-luciferase neutralization assay. (**A**) Live virus assay for Wuhan wild type WT vs. Delta for all ages and (**B**) live virus assay for WT vs. Delta for young adults and the elderly. (**C**) Pseudovirus assay for WT vs. BA.1 for all ages and (**D**) Pseudovirus assay WT vs. BA.1 for young adults and the elderly. (**E**) Pseudovirus assay for WT vs. BA.2 for all ages and (**F**) Pseudovirus assay WT vs. BA.2 for young adults and the elderly. The 50% viral-neutralizing antibody geometric mean titers (GMT, 95% CI) were measured, VNT_50_ for live virus and pVNT_50_. Statistical analysis was performed by the Student’s t-test (ns, *p*>0.05; ****, *p*<0.0001). No significant difference is notable between the two age groups in neutralization effect against WT, Delta, BA.1, and BA.2.

As to Omicron BA.1 and BA.2 subvariants when they sequentially dominated the pandemic scene, neutralization effects were measured by using pseudovirus for WT and Omicron subvariants. UB-612 booster elicited high neutralizing titers against WT at pVNT_50_ of 6,245; versus that against BA.1 at 1,196, representing a 5.2-fold reduction (**Figure 2C**); and versus against BA.2 at pVNT50 of 985, representing a 6.3-fold reduction (**Figure 2E**). There was no significant age-dependent booster-induced neutralization effect between young adults (18-65 yrs.) and the elderly (65-85 years) with respect to either anti-WT or anti-Omicron pVNT_50_ level (**Figure 2, D and F**). Both age groups showed a 5.0- to 7.6-fold reduction for anti-BA.2 relative to anti-WT. By all accounts compared with BA.1, booster vaccination exhibits only a minor 1.2-fold lower neutralizing activity against BA.2.

### Neutralizing antibodies against WT, Omicron BA.1, BA.2, and BA.5

On the occasion when the pandemic is being dominated by the ongoing BA.5 subvariant, we tested samples from 12 participants from the two groups (18-65 years, n = 7; 65-85 years, n = 5), with neutralization titers measured using pseudovirus. UB-612 booster elicited neutralizing titers of pVNT_50_ against WT/BA.1/BA.2/BA.5 at 11,167/2,314/1,890/854 (**Figure 3A**), representing a 4.8-/5.9-/13-fold reduction, respectively, relative to the anti-WT level. By comparison with BA.2, booster vaccination exhibits only a modest 2.0-fold lower neutralizing activity against BA.5. There was no statistically significant difference age-dependent neutralization effect between young adults and the elderly within each of anti-WT/-BA.1/-BA.2/-BA.5 pVNT_50_ levels (**Figure 3, B-D**). By all accounts compared with BA.2, booster vaccination exhibits only a modest 2-fold lower neutralizing activity against BA.5.

**Figure 3.**
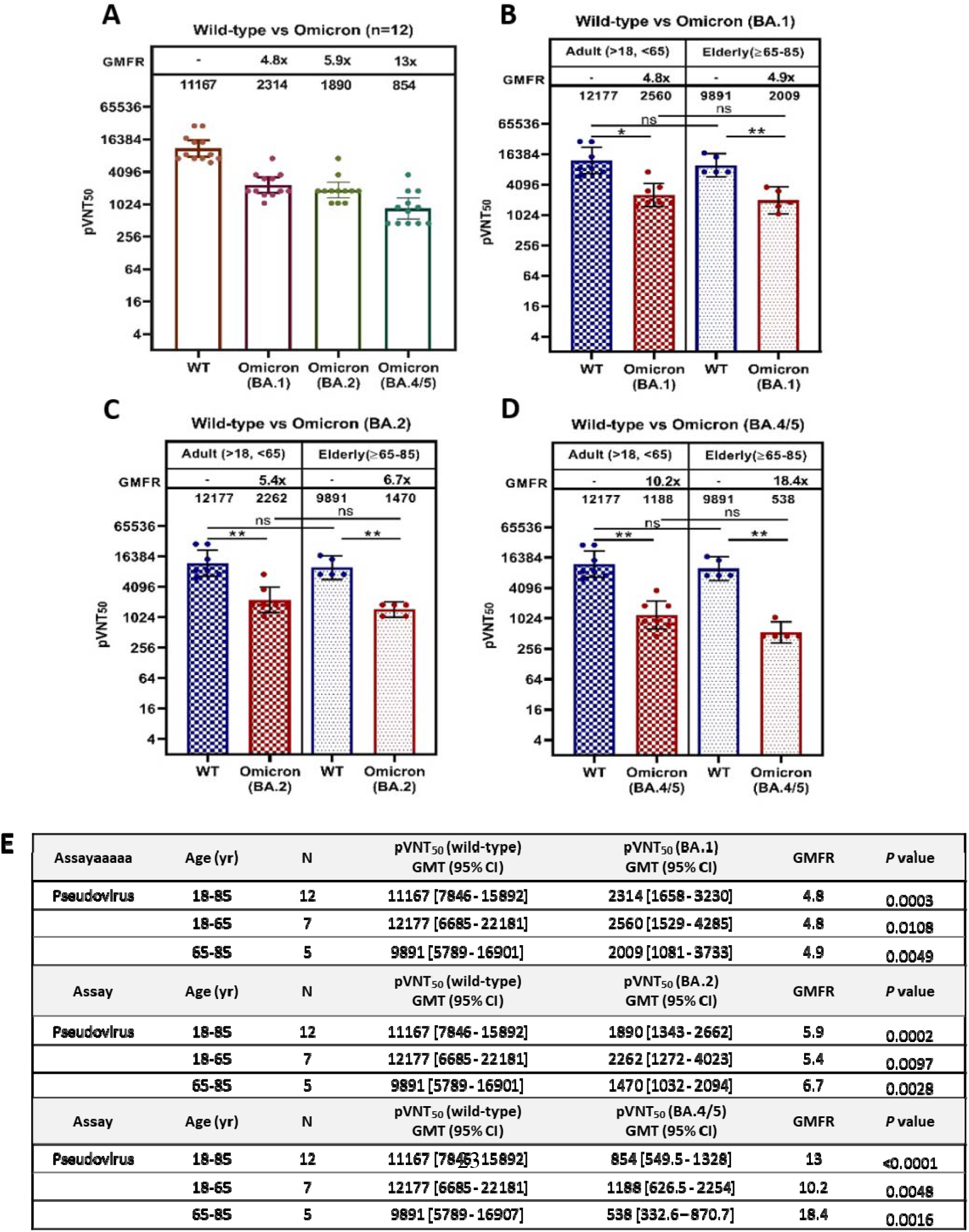
Comparative viral-neutralization effects against wild type strain, Omicron BA.1, BA.2, and BA.5 variants. At the SARS-CoV-2 pandemic when the Omicron BA.5 variant dominates, viral-neutralizing titers against wild-type, Omicron BA.1, BA.2, and BA.5 variants were measured using pseudovirus assay for comparison. Serum samples from 12 participants (n = 7 for 18-65 years; n = 5 for 65-85 years) collected at 14 days post-booster were subjected to pseudovirus-luciferase neutralization assay. (**A**) Wuhan wild type WT, BA.1, BA.2, and BA.5 for study participants of all ages, (**B**) WT vs. BA.1 for young adults and the elderly, (**C**) WT vs. BA.2 for young adults and the elderly, and (**D**) WT vs. BA.5 for young adults and the elderly. The 50% viral-neutralizing antibody geometric mean titers (GMT, 95% CI) were measured, pVNT_50_. Statistical analysis was performed by the Student’s t-test (ns, *p*>0.05; ****, *p*<0.0001). No significant difference is notable between the two age groups in neutralization effect against WT, BA.1, BA.2, and BA.5.

### Potent, durable and correlative viral-neutralization and ACE2:RBD_WT_ binding inhibition

We explore the correlation between viral-neutralizing activity (VNT_50_) and receptor binding inhibition (ACE2:RBD_**WT**_) using immune sera from 87 participants available on Day 1 (pre-dose), Day 57 (28 days post-2^nd^ dose), Day 220 (pre-boosting, 6 to 8 months post-2^nd^ dose), and Day 234 (14 days post-booster) which showed a high post-booster VNT_50_ titer of 738 (**Figure 4A**), a 17-fold increase over the pre-boosting (titer 44) and a 7-fold increase over the levels of both Day 57 (titer 104) and the human convalescent sera, HCS (titer 102).

**Figure 4.**
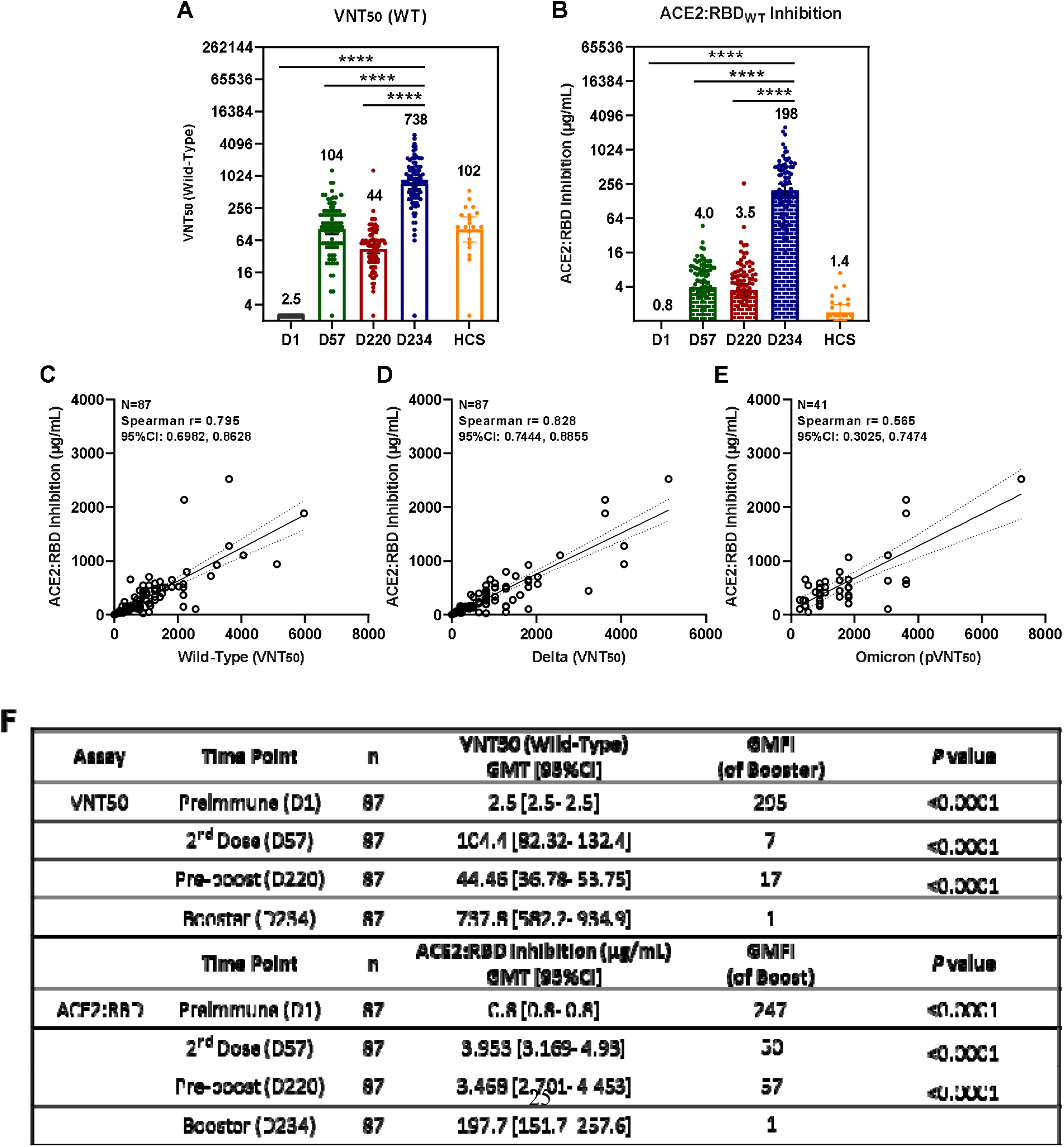
Functional correlations between ACE2:RBD_WT_ binding inhibition and viral-neutralization against Delta and Omicron BA.1. Of 871 participants enrolled in the phase II primary 2-dose series and grouped for Immunogenicity investigation, serum samples from 87 participants who had received a booster 3^rd^-dose of 100 μg UB-612 were collected at Day 1 (pre-dose 1), Day 57 (28 days post-dose 2), Day 220 (pre-booster between Days 197 to 242,), Day 234 (14 days post-booster between Days 211 to Day 256). HCS from 20 SARS-CoV-2 infected individuals were also included for comparative testing by two functional assays: **(A)** viral-neutralizing titer (VNT_50_) against live wild-type Wuhan strain (WT) by CPE method; and **(B)** the antibody concentration calibrated with an internal standard for ACE2:RBD_WT_ binding inhibition by ELISA. The correlations are explored between the two function assays, i.e., ACE2:RBD_WT_ binding inhibition ELISA and the viral-neutralizing titers against the live virus (VNT_50_ for original wild-type and Delta strains) or the psuedovirus (pVNT_50_ for Omicron BA.1 strain). The RBD_WT_ stands for the RBD binding protein domain bearing amino acid sequence of the original SARS-CoV-2 wild-type (WT) Wuhan strain. The correlations were explored for **(C)** ACE2:RBD_WT_ inhibition vs. anti-WT VNT_50_, **(D)** ACE2:RBD_WT_ inhibition vs. anti-Delta VNT_50_, and **(E)** ACE2:RBD_WT_ inhibition vs. anti-Omicron pVNT_50_. The correlation coefficients were evaluated by Spearman r with 95% CI. Statistical analysis was performed with the Student’s t-test (ns, *p*>0.05; ***, *p*<0.001; ****, *p*<0.0001). **(F)** Summary of geometric mean titer (GMT) and 95% CI are presented for plots as shown in Figures (**A**) and (**B**).

In addition, a pronounced post-booster functional antibody-mediated inhibition of ACE2:RBD_**WT**_ binding inhibition was also observed at a high titer (expressed in standard-calibrated antibody concentration) of 198 μg/mL (**Figure 4B**), a ∼57-fold increase over both the pre-boosting titer of 3.5 μg/mL and the Day 57 of 3.5 μg/mL, and a profound 140-fold over the HCS titer of 1.4 μg/mL. With increased monitoring time, the results again demonstrated that UB-612 induces a durable neutralizing antibody titer level, observed between Day 57 (post-2^nd^ dose) vs. Day 220 (pre-boosting), which represents a 42% retainment for VNT_50_ (titer, 104 vs. 44) against live WT virus (**Figure 4A**) and a 88% retainment for ACE2:RBD_**WT**_ (μg/mL, 4.0 vs. 3.5) binding inhibition (**Figure 4B**).

The inhibition of ACE2:RBD_**WT**_ binding on ELISA correlates well with anti-WT (**Figure 4C**) and anti-Delta VNT_50_ (**Figure 4D**) findings, both showed a similar high correlative Spearman’s rank correlation coefficients (r = 0.795 and 0.828, respectively). A lesser but significant correlation was also observed for ACE2:RBD_WT_ binding inhibition and anti-Omicron BA.1 pVNT_50_, with a Spearman’s correlation coefficient of r = 0.565 (**Figure 4E**).

## DISCUSSION

UB-612 booster, proven safe and well tolerated without concerns of SAEs (**Supplemental Figure 2**), induces potent memory T cell immunity (**Figure 1**) that synergizes recalled B cell immunity with striking cross-neutralizing antibodies against WT, Delta and Omicrons (**Figures 2 and 3**). Of notable clinical interest, the booster uplifts a low neutralizing antibody titer in the elderly (43) to a high level close to that in young adults regardless of viral mutant status (**Figures 2 and 3**). In addition, blockade of S1-RBD binding to ACE2 receptor well correlates with viral neutralization (**Figure 4**). Thus, the UB-612 vaccine platform, due to broadly recognizing conserved Th/CTL epitopes on Spike and non-Spike proteins, can maintain a target plasticity without much mutational distortion. The present report reveals five salient findings.

Firstly, on the magnitude of pseudovirus-neutralizing titer (pVNT_50_/ID_50_) against Omicrons, UB-612 booster offers a potential competitive edge (**Table 2A**). While the rank order of pVNT_50_ against Omicrons BA.1/BA.2/BA.5 trends downward (**Figure 3A**) and UB-612 booster combats the most contagious BA.5 with a 13-fold reduction relative to anti-WT titer, its anti-BA.5 pVNT_50_ titer of 854 represents a substantially high neutralizing activity. In contrast, an anti-BA.5 pVNT_50_ titer of 582 was reported for NVX-CoV2373 (44), 378 for mRNA-1273 (7), 360 for BNT162b2 (2, 3), 75 for CoronaVac (6) and 43 for AZD1222 (2) (**Table 2A**). All available anti-BA.5 pVNT_50_ values account for approximately an 8.4- to 21-fold reduction. There is a stark vaccine platform-dependent difference in the anti-BA.5 potency.

**Table 2.** Viral-neutralizing antibody titers against SARS-CoV-2 wild-type (WT) and Omicron BA.1, BA.2 and BA.5 subvariants upon homologous boosting.

**(A) Pseudotyped virus neutralization assay (pVNT<sub>50</sub> and ID<sub>50</sub>)**
| Vaccines<sup>a</sup><br>(Homo-booster)<sup>a</sup> | Participants<br>(No./Reference) | NeuAb assay<br>(Method/Unit) | WT<br>(GMT)<sup>b</sup> | Omicrons<br>BA.1/BA.2/BA.5<br>(GMT)<sup>b</sup> | WT/Omicrons<br>BA.1/BA.2/BA.5<br>(GMFR) |
| --- | --- | --- | --- | --- | --- |
| UB-612 (Ph-1) | (n = 45)/(ref. 43) | PNA/pVNT<sub>50</sub> | 12,778 | 2,325/ND/ND | 5.5/ND/ND |
| UB-612 (Ph-2)<sup>c</sup> | (n = 41)/(present) | PNA/pVNT<sub>50</sub> | 6,254 | 1,196/985/ND | 5.2/6.3/ND |
| UB-612 (Ph-2)<sup>c</sup> | (n = 12)/(present) | PNA/pVNT<sub>50</sub> | 11,167 | 2,314/1,890/854 | 4.8/5.9/13.0 |
| BNT162b2 | (n = 24)/(ref. 20) | PNA/pVNT<sub>50</sub> | 6,539 | 1066/776/ND | 6.1/8.4/ND |
| BNT162b2 | (n = 19)/(ref. 2) | PNA/pVNT<sub>50</sub> | 4,122 | 1,116/1,113/360 | 3.7/3.7/11.5 |
| BNT162b2 | (n = 27)/(ref. 3) | PNA/ID<sub>50</sub> | 5783 | 900/829/275 | 6.4/7.0/21.0 |
| NVX-CoV2373 | (n = 48)/(ref. 44) | PNA/ID<sub>50</sub> | 10,862 | 1,197/ND/582 | 9.1/ND/18.7 |
| mRNA-1273 | (n = 16)/(ref. 7) | PNA/ID<sub>50</sub> | 4,679 | 945/780/378 | 5.0/5.9/12.4 |
| MVC-COV1901 | (n = 15)/(Ref. 45) | PNA/ID<sub>50</sub> | 1,280-640 | 160-80/ND/ND | 8.7/ND/ND |
| CoronaVac | (n = 40)/(Ref. 6) | PNA/pVNT<sub>50</sub> | 632 | 122/122/75 | 5.2/5.2/8.4 |
| AZD1222 | (n = 41)/(Ref. 2) | PNA/pVNT<sub>50</sub> | 516 | 89/76/43 | 5.8/6.8/12 |
| BBIBIP-CorV | (n = 75)/(ref. 46) | PNA/pVNT<sub>50</sub> | 295 | 15/ND/ND | 20.1/ND/ND |

**Abbreviation:** PNA = pseudotyped virus neutralization assay; GMT = geometric mean titer; GMFR = geometric mean fold reduction relative to WT; WT = wild type strain of SARS-CoV-2; Omicrons = Omicron subvariants BA.1/BA.2/BA.5; ND = not determined. pVNT<sub>50</sub> & ID<sub>50</sub> = 50% neutralization GMT by pseudovirus assay
<sup>a</sup> Vaccines reported of homologous booster (third dose) vaccination. <sup>b</sup> GMTs against WT measured at 14 or 28 days post-booster third dose. <sup>c</sup> UB-612 - Pseudovirus assays conducted with sera of subset participants (Phase-2 booster extension study) when Omicron infection were dominated sequentially by BA.2 and BA.5 subvariant.

**(B) Live virus neutralization assay (VNT<sub>50</sub>, PRNT<sub>50</sub>, and FRNT<sub>50</sub>)**
| Vaccines<sup>a</sup><br>(Homo-booster)<sup>a</sup> | Participants<br>(No./Reference) | NeuAb assay<br>(Method/Unit) | WT<br>(GMT)<sup>b</sup> | Omicrons<br>BA.1/BA.2<br>(GMT)<sup>b</sup> | WT/Omicrons<br>BA.1/BA.2<br>(GMFR) |
| --- | --- | --- | --- | --- | --- |
| UB-612 | (n = 15)/(ref. 47) | MNA/VNT<sub>50</sub> | 6,159 | 670/485 | 9.2/12.7 |
| mRNA-1273 | (n = 20)/(ref. 7) | FRNT/ID<sub>50</sub> | 1659 | 81.0/ND | 20.5/ND |
| BNT162b2 | (n = 20)/(ref. 7) | FRNT/ID<sub>50</sub> | 640 | 46.2/ND | 13.3/ND |
| BNT162b2 | (n = 30)/(ref. 29) | PRNT/VNT<sub>50</sub> | 673 | 106/ND | 6.3/ND |
| AZD1222 | (n = 41)/(Ref. 48) | FRNT/FRNT<sub>50</sub> | 723 | 57.0/ND | 12.7/ND |

**Abbreviation:** MNA = Microneutralization assay; PRNT = plaque reduction neutralization test; FRNT = focus reduction neutralization test; GMT = geometric mean titer; GMFR = geometric mean fold reduction relative to WT; WT = wild type strain of SARS-CoV-2; Omicrons = Omicron subvariants BA.1/BA.2; ND = not determined. pVNT<sub>50</sub> & ID<sub>50</sub> = 50% neutralization GMT by live virus assay; VNT<sub>50</sub>, ID<sub>50</sub> & FRNT<sub>50</sub> = 50% neutralization GMT by live virus assay.
<sup>a</sup> Vaccines reported of homologous booster (third dose) vaccination. <sup>b</sup> GMTs against WT measured at 14 or 28 days post-booster third dose.

High anti-WT/anti-BA.1 pVNT_50_ titers of 12,778/2,352 for UB-612 were first observed in our Phase-1 booster vaccination study (43) (**Supplemental Figure 4B**). As to “anti-WT vs. anti-BA.1 vs. anti-BA.2,” UB-612 booster exhibits a combined pVNT_50_ value profile of 6254-12,778 vs. 1196-2325 vs. 985-1819” (**Table 2A**), which are comparable to the respective counterparts reported for NVX-CoV2373 (44), mRNA-1273 (7), BNT162b2 (2), yet far greater than for MVC-COV1901 (45), CoronaVac (6), AZD1222 (2), and BBIP-CorV (46). All listed anti-BA.1/BA.2 pVNT_50_ values account for approximately a ∼3.7- to 20-fold reduction; and, relative to anti-BA.1/-BA.2, the overall anti-BA.5 pVNT_50_ is estimated to be within 2- to 3-fold reduction. Altogether, UB-612 booster performs at least on a par with or bear a competitive edge over other vaccine platforms in terms of pseudovirus-neutralizing pVNT_50_, against Omicron BA.1/BA.2/BA.5. In comparative view of between-vaccine platforms, the magnitude of viral-neutralizing strength would matter much more than a sheer GMFR factor.

Secondly, on the level of live virus-neutralizing titer (VNT_50_/ID_50_/FRNT_50_) against Omicron BA.1 and BA.2 (**Table 2B**), UB-612 booster excels over other vaccine platforms. UB-612 booster elicits an anti-WT/anti-BA.1 titer profile of 6,159/670 (47), in contrast with 1699/81.0 for mRNA-1273 (7), 640-673/46.2-106 for BNT162b2 (7, 29), and 723/57 for AZD1222 (48). UB-612’s anti-BA.1 titer of 670 far exceeds other vaccines by ∼6- to 12-fold higher. Of note, other vaccine platforms exhibit a low anti-BA.1 level at peak response (28 days post-booster), which could wane rapidly so that a fourth-dose jab (the 2^nd^ booster) has been proposed to compensate their dwindling neutralizing antibodies.

In addition, UB-612 booster presents a substantially high anti-BA.2 live-virus titer VNT_50_ at 485, which is far greater than the anti-BA.1 titers observed with other vaccines. In light of the true measure for neutralizing activity, the live virus assay would reflect better than by pseudovirus assay, as the former stands for the combined anti-viral activity against both Spike and non-Spike, while the latter against the Spike only.

Similar high neutralizing titers against live WT/Delta virus (VNT_50_) have been noted earlier for UB-612 in the Phase-1 booster study (43). By contrast, the post-booster VNT_50_ values against WT/Delta range from the low-end 122/54 (CoronaVac) to the high-end 3,992/2358 (UB-612) **(Supplemental Table 1)**. In the present Phase-2 booster study, UB-612 reproduces a high anti-Delta VNT_50_ at 1,282, only a 1.3-fold lower than the anti-WT live virus (**Figure 2A**).

Collectively, UB-612 booster performs at least on a par with or bear a competitive edge over other vaccine platforms in viral-neutralization potency, either pseudovirus or live virus, against Delta and Omicron BA.1, BA.2, and potentially the currently dominating BA.5.

Thirdly, UB-612 booster uplifts a low viral-neutralizing titer generally associated with the elderly to a level approximately the same as that in the young adults. No significant age-dependent neutralization effect is evident between young adults and the elderly with respect to humoral immune responses against WT/Delta/BA.1/BA.2/BA.5 (**Figure 2, B, D, and F; Figure 3, B-D**). This is of high clinical significance as elderly people, due to a decline in pathogen immunity, do not respond to immune challenge as robustly as the young adults and so come with a reduction in vaccine efficacy (49). UB-612 as a primer or booster thus has a potential benefit not only for the elderly but also for immunocompromised people in general.

Fourthly, the strong blockade of ACE2:RBD_WT_ interaction (**Figure 4B**) correlates well with viral-neutralizing VNT_50_ (live virus WT and Delta) and pVNT_50_ (pseudovirus BA.1) **(Figure 4, C-E)**. The positive functional correlation infers a substantial clinical efficacy against COVID-19. Indeed, using models of S protein binding activities (50) and neutralizing antibodies (51), the clinical efficacy of 2-dose primary immunization of UB-612 is predicted to be 70-80% and a booster vaccination may lead to 95% efficacy against symptomatic COVID-19 caused by the ancestral Wuhan strain (47). The clinical efficacy protecting against infection of circulating subvariants including the dominant B5 would await outcome of an ongoing Phase-3 trial that compares UB-612 with authorized vaccines under homologous and heterologous boosting [ClinicalTrials.gov ID: NCT05293665].

The pronounced, broadly-neutralizing profiles illustrate one unique feature of UB-612, i.e., the serum neutralizing antibodies are directed solely at the critical receptor binding domain (RBD) that reacts with ACE2. In contrast with the currently authorized full-length S protein-based vaccine platforms, UB-612’s RBD-only design leaves little room in non-conserved sites of S protein for viral mutation to occur and so may result in an immunity with less immune resistance.

Thus, booster vaccination can prompt recall of high levels of parallel anti-WT neutralizing VNT_50_ (**Figure 4A**) and RBD-ACE2 binding inhibition antibodies (**Figure 4B)**, and both functional events are durable over Day 57 and Day 220 with a substantial 42%/88% retainment at 6 months or longer after the second shot. This is consistent with the long-lasting anti-WT VNT_50_ effect with a half-life of 187 days observed in the phase I primary series, in which a ∼50% retainment was observed at 6 months relative to the peak response (43).

Further, the finding that the UB-612 induced a 140-fold higher increase in blocking the RBD:ACE2 interaction than by human convalescent sera (HCS) (**Figure 4B**) suggests that most of the antibodies in HCS may bind allosterically to the viral spike (N- or C-terminal domain of the S) rather than orthosterically to the RBD sites, which may include non-neutralizing anti-S antibodies to cause unintended side effects or ADE event. This warrants further investigation including sera from re-infections and breakthrough infections from all vaccine platforms.

Fifthly, UB-612 booster induces potent, durable Th1-oriented (IFN-γ^+^-) responses (peak/pre-boost/post-boost SFU/10^6^ PBMCs, 374/261/444) along with robust presence of cytotoxic CD8^+^ T cells (peak/pre-boost/post-boost CD107a^+^-Granzyme B^+^, 3.6%/1.8%/1.8%) (**Figure 1**). Vaccines designed to produce a strong systemic T cell response targeting conserved nonmutable epitopes may prevent immune escape and protect against current and future viral variants that causes COVID-19 (52, 53). Along the same vein of promoting T cell immunity, UB-612 armed with pool of sequenced-conserved Th/CTL epitope peptides (S2×3, M, and N) (**Table 1B**) has demonstrated to elicit a striking, durable Th1-predominant IFN-γ^+^ T cell response in Phase-1 primary vaccine series that peaked at 254 SFU/10^6^ and persisted with a ∼50% retainment over 6 months post-2^nd^ dose (121 SFU/10^6^ cells) (43).

The Phase-1 booster-recalled T cell immunity is consistent with an even higher 70% sustaining T cell immunity (peak 374 vs. pre-boost 261 SFU/10^6^ cells) in the present Phase-2 primary series that surges to a 444 SFU/10^6^ cells two weeks post-booster (**Figure 1, A and D**), which leads to a pronounced, durable cytotoxic (CD107a^+^-Granzyme B^+^) activity of CD8^+^ T lymphocytes (CTL) with high frequency levels (1.8%-3.6%) (**Figure 1, C and F**).

UB-612 booster appears to trigger far greater T cell responses than those produced by the current Spike-only mRNA (BNT162b2) and adeno-vectored (ChAdOx1) vaccines (54): e.g., the pre-boost/post-boost level of SFU/10^6^ cells (related to Delta strain) under homologous boosting for the 3-dose of ChAd/ChAd/ChAd were 38/45 and that of BNT/BNT/BNT were 28/82; and those under heterologous boosting were 42/123 for ChAd/ChAd/BNT and 36/108 for BNT/BNT/ChAd. For those currently licensed COVID vaccines, it is worthy to note that the fourth vaccine jab (the 2^nd^ booster) does not increase T cell response (55): e.g., the 28 days post-3^rd^ dose/pre-4^th^ dose/14 days post-booster level of SFU/10^6^ cells (related to Delta strain) for the 4-dose of ChAd/ChAd/BNT/BNT were 133/19/108 and that of BNT/BNT/BNT/BNT were 62/14/80.

The lackluster booster-recalled T cell immunity seen with mRNA and adeno-vectored vaccines (54, 55) may reflect the dwindling, weakened B cell humoral responses and clinical efficacy. A booster 3^rd^-dose of mRNA vaccines could compensate the waning immunity and reduce rates of hospitalization and severe disease, yet less effective in protection against mild and asymptomatic infections (25-30). At the time of Omicron BA.1 on the rise, vaccine effectiveness was seen reduced after booster third dose of mRNA vaccines in protection against COVID symptoms (45% at 10 weeks) (56) and hospital admission (55% at 12 weeks) (57).

In two retrospective large cohort studies, the elderly (aged ≧60) receiving the fourth dose of BNT162b2 (2^nd^ booster) while BA.2 infection was dominant also showed a modest and transient efficacy against severe disease (∼60-75% protection, relative to the 1^st^ booster third dose) (58, 59), and the effectiveness against infection completely wanes after 8 weeks (58).

Breakthrough infection could occur after the fourth dose (31, 32), in particular amid the circulation of the dominant Omicron BA.5. The booster-compensated protection effectiveness offered by mRNA vaccines could be blunted soon upon boosting. Incessant, short-interval boosting with current mRNA vaccines could result in dwindling and weakened immune responses against Omicrons (60), for mechanisms remained to be elucidated.

While a substitute of the fourth dose with mRNA-1273 can elevate T cell response to a level of ∼240 SFU/10^6^ cells (55), the increased response level appears to be lower than those by UB-612 at 261 SFU/10^6^ cells pre-3^rd^-dose boosting and at 444 SFU/10^6^ cells 14 days post-booster (third dose) (**Figure 1**). Presumably, UB-612 vaccine designed to target multiple conserved epitopes on both Spike and non-Spike proteins could have underpinned the base for a fuller T cell immunity.

The potential clinical significance of a striking T-cell immunity elicited by UB-612 vaccine platform is supported by the development of a plain T-cell vaccine (CoVac-1) containing a six-peptide backbone that, as a T-cell booster, triggered dramatic multifunctional CD4 and CD8 T-cell responses (61), which showed benefits to B-cell deficient, immunocompromised patients who could not mount B-cell antibody responses. The facts that potent memory CD4 and CD8 T cell memory can protect against SARS-CoV-2 infection in the absence of immune neutralizing antibodies (52, 53, 61) raises concerns over the fact that humoral antibody response has long been used as a sole bridging metric of protective immunity (62), which lacks full understanding of human post-vaccination immunity as antibody response is generally shorter-lived than virus-reactive T cells (63-65).

Further, the SARS-CoV-2’s non-Spike structure E, M and N proteins are the regions critically involved in the host cell interferon response and T-cell memory (36-38). These structural proteins of virus’ main body when picked up by Antigen Presenting Cells (APC) and presented as viral peptides would fall beyond recognition by the currently authorized vaccines that are based on the outer spike protein only. Th1 cells help to stimulate B cells to make antibodies, and they can morph as well into memory helper CD4^+^ and cytotoxic CD8^+^ T cells to provide a long-lasting immune response (66). UB-612’s booster-enhanced broader, durable B and T cell immunity may make Omicron evasion less likely as the booster vaccination could behave closer to the breadth of natural, infection-induced immunity.

While neutralizing antibodies can block ACE2:RBD interaction and viral infection, the non-Spike protein cross-reactive memory T cell immunity is essential for effective long-term prevention against infection and should be recognized as a measure for the long-term vaccine success (67-74). The role of T cells, in particular the recognition against non-Spike targets and the associated T cell responses, has long been underestimated, overlooked from the outset of COVID vaccine development. To what extent that vaccine booster-induced memory T cell immunity would contribute to vaccine effectiveness in the clinic against COVID-19 infection of any degree, as a leading actor or a supportive cast, has become a research subject of major clinical interest (75).

Of additional clinical interest with strong T cell immunity is its function of viral clearance. Persistent SARS-CoV-2 infections can contribute to long-haul COVID as residual viable SARS-CoV-2 particles, viral replication, viral RNA and viral spike protein antigens could sustain in tissues of the convalescents (76-78). Spike S1 protein could persist in monocytes that could cause inflammation and lung tissue damage (79). As long-haul COVID is found to be associated with a decline in IFNγ-producing CD8^+^ T cell (80), enhancing T cell immunity for clearance of residual systemic infection (sustained viral reservoirs) could be a sensible strategy for prevention of long-haul COVID.

Facing the dwindling vaccine effectiveness and emergence of viral variants with higher infectivity and immune evasion, development of composition-updated vaccines (33, 34) or universal coronavirus vaccines (35) has been strongly advocated. To meet an urgent need and for a long-term sense, one would look beyond the practice of frequent short-interval booster jabs and resist clinging to use of variant-specific (e.g., omicron-updated) vaccines. In fact, the recent mRNA bivalent vaccine (original Spike plus Omicron BA.1 Spike) as the fourth dose (second booster) was found to result in only 1.2- to 1.8-fold higher neutralizing antibody titer (pVNT_50_) against BA.1 when compared to the ancestral wild-type strain (81).

A pragmatic approach to curbing ever-emergent new mutants would be “universal (pan-Sarbecovirus) vaccines” targeting conserved nonmutable epitopes on coronavirus. In that sense, a shift of Spike-only vaccine design to a paradigm by targeting conserved epitopes on both Spike and non-Spike proteins would be a workable option. And, to be competent for next-generation vaccines, conserved regions on non-Spike proteins (membrane and nuleocapsid) to serve as immunogens may also contribute to the development of pan-betacoronavirus vaccines (82).

By incorporation of five sequence-conserved Th/CTL epitope peptides and a sixth idealized universal Th peptide which serves as catalyst in T cell activation, the UB-612-induced T cell immunity may enhance the clearance of the virally infected cells regardless of Omicrons or future mutants, as their mutation sites are not to overlap any of the amino acid residues on the precision-designed S2, N, and M epitope peptides that are highly conserved (or rarely mutate) across all VoCs (83) (**Table 1B**). The viral proteins on Omicron that are substantially conserved have been reported as strong T-cell activators and induce long-lasting T-cell responses (84), which can synergize with B-cell memory for enhanced immunity.

In summary, we have simultaneously characterized the booster-enhanced B- and T-cell immunity in a large Phase-2 study, demonstrating UB-612 can elicit a fuller T cell immunity that comprehensively recognize Spike (S1-RBD and S2) and non-Spike structure N and M proteins, which seeds the potential for viral clearance upon infection; and the induced B cell responses would broadly neutralize all VoCs regardless of varying mutational epitope locations. Our UB-612 multitope-vaccine with recent EUA approvals in two Asian countries, may serve as a universal vaccine primer and booster to ward off all VoCs and future mutants, for which a US-FDA approved CEPI supported large-scale phase III trial has also been underway to further prove the concept of protection effectiveness.

## Data Availability

All data produced in the present work are contained in the manuscript.

## AUTHOR CONTRIBUTIONS

CYW, KPH, WJP, HL and YHS were responsible for vaccine development including vaccine formulation design, protocol design and implementation of the clinical studies, acquisition and interpretation of the clinical data. MSW and YTY were responsible for assay development and validation, laboratory testing and data collection, and preparation of respective reports. CYW, WJP, YHH, HL, and BSK had full access to and verified all the data in the study and take responsibility for the integrity and accuracy of the data analysis. BSK and CYW drafted and prepared the manuscript. Drs. Chuwan-chuen King and Carl V. Hanson provided critical review of the manuscript. All authors reviewed and approved the final version of the manuscript. CYW had final responsibility for the decision to submit for publication.

## DATA SHARING

The study protocols are provided in the Supplemental Appendices. Individual participant data will be made available when the trial is complete with data to be shared through a secure online platform.

## DECLARATION OF INTERESTS

CYW is co-founder and board member of UBI, United BioPharma, and UBI Asia, and named as an inventor on several patent applications filed covering COVID vaccine development. HKK and WJP are also named as co-inventors on related patent applications. CYW, WJP, BSK, HL, YHH, MSW, YTY, and PYC are employees within the UBI group.

## ACKNOWLEDGEMENTS

The study was funded by UBI Asia (study sponsor) and the Taiwan Centers for Disease Control, Ministry of Health and Welfare. The sponsor co-designed the trial and coordinated interactions with contract Clinical Research Organization (CRO) StatPlus staff and regulatory authorities. The CRO took charge of trial operation to meet the required standards of the International Council for Harmonization of Technical Requirements for Pharmaceuticals for Human Use and Good Clinical Practice guidelines. The Independent Data Monitoring Committee (IDMC) oversaw the safety data and gave recommendations to the sponsor. The interim analysis was done by the CRO StatPlus. We thank all the trial participants for their dedication to these trials; the investigation staff at China Medical University Hospital, Taipei

Medical University Hospital, Far Eastern Memorial Hospital, National Cheng Kung University Hospital, Linkou Chang Gung Memorial Hospital, Kaohsiung Chang Gung Memorial Hospital, Kaohsiung Medical University Hospital, Tri-Service General Hospital, Taipei Veterans General Hospital, Kaohsiung Veterans General Hospital, Changhua Christian Hospital and Taichung Veterans General Hospital for their involvement in conducting the trial; and members of the IDMC for their dedication and guidance.

Special thanks are also extended to the clinical associates from StatPlus, Inc and UBI Asia; the CMC task forces from UBI Asia, United BioPharma, Inc. and UBI Pharma, Inc; team members at Institute of Biomedical Sciences, Academia Sinica for the live virus neutralization assay; and team members at the RNAi Core Facility, Academia Sinica for the pseudovirus neutralization assay. All health convalescent sera were supplied by Biobank at the National Health Research Institutes (NHRI), Taiwan.

Finally, thank colleagues from UBI Asia Hui-Kai Kuo, Wan-Yu Tsai, Han-Chen Chiu, Kuo-Liang Hou, and Jennifer Cheng for their technical support and data collection. Special administrative support by Jalon Tai, Liang Kai Huang, Peter Hu and Fran Volz from the UBI group are also acknowledged with gratitude.

## Notes

### Competing Interest Statement

The authors have declared no competing interest.

### Clinical Trial

NCT04773067

### Author Declarations

Ethics Committee of China Medical University Hospital gave ethical approval for this work Ethics Committee of Taiwan Medical University gave ethical approval for this work

