## Supplemental Information for "UB-612 Multitope Vaccine Targeting SARS-CoV-2 Spike and Non-Spike Proteins Provides Broad and Durable Immune Responses"

Chang Yi Wang, on assignment at UBI Asia, Hsinchu, Taiwan

**Key words:** UB-612, Multitope Universal Vaccine, booster vaccination, SARS-CoV-2, Sarbecovirus, and Variants of Concern.

**Supplemental Information**

**Supplemental Figures**

Figure S2. Incidence of adverse effects in the Phase-2 primary 2-dose and extended booster third-dose series……………………………………………………………………….4

Figure S3. Phase-2 immunogenicity overview (antigenic and functional) on homologous boosting……………………………………………………………………………….5

Figure S4. Viral-neutralizing titers against live SARS-CoV-2 wild type (Wuhan) and Delta

variant (VNT_50_), and pseudo SARS-CoV-2 wild type (Wuhan) and Omicron

variant (pVNT_50_) after the booster third-dose in the Phase-1 trial……………………6

**Supplemental Tables**

Table S1. Comparison of post-booster viral-neutralizing antibody titers against SARS-CoV-2

wild-type (WT) and Delta variant by vaccines from different platforms………………7

**Supplemental Methods**

￭ Viral-neutralizing antibody titers against SARS-CoV-2 wild-type and variants by CPE-based live virus neutralization assay…………………………………………………………………..8

￭ Neutralizing titers against Omicron BA.1/BA.2/BA.5 by pseudovirus assay...………...............9

￭ [Inhibition of RBDWT binding to ACE2 by ELISA…………………………………………...10](#_Toc95429259)

￭ Anti-S1-RBDw_T_ binding IgG antibody by ELISA…………………………………………….11

￭ T cell responses by ELISPOT………………………………………………………………….11

￭ Intracellular cytokine staining…………………………………………………………………11

￭ Statistics………………………………………………………………………………………..12

**Supplemental Appendices** (available upon request)……………………………………………13

Appendix 1: Phase 2 study V-205 protocol

Appendix 2: Phase 2 study V-205 IRB approval letter 1

Phase 2 study V-205 IRB approval letter 2

Phase 2 study V-205 IRB approval letter 3

Appendix 3: Phase 2 study V-205 Informed Consent Form (ICF)

Appendix 4: CONSORT checklist

**Supplemental Figures**

**Figure** **S1. Flow** **of UB-612 phase II primary 2-dose series with extension booster**

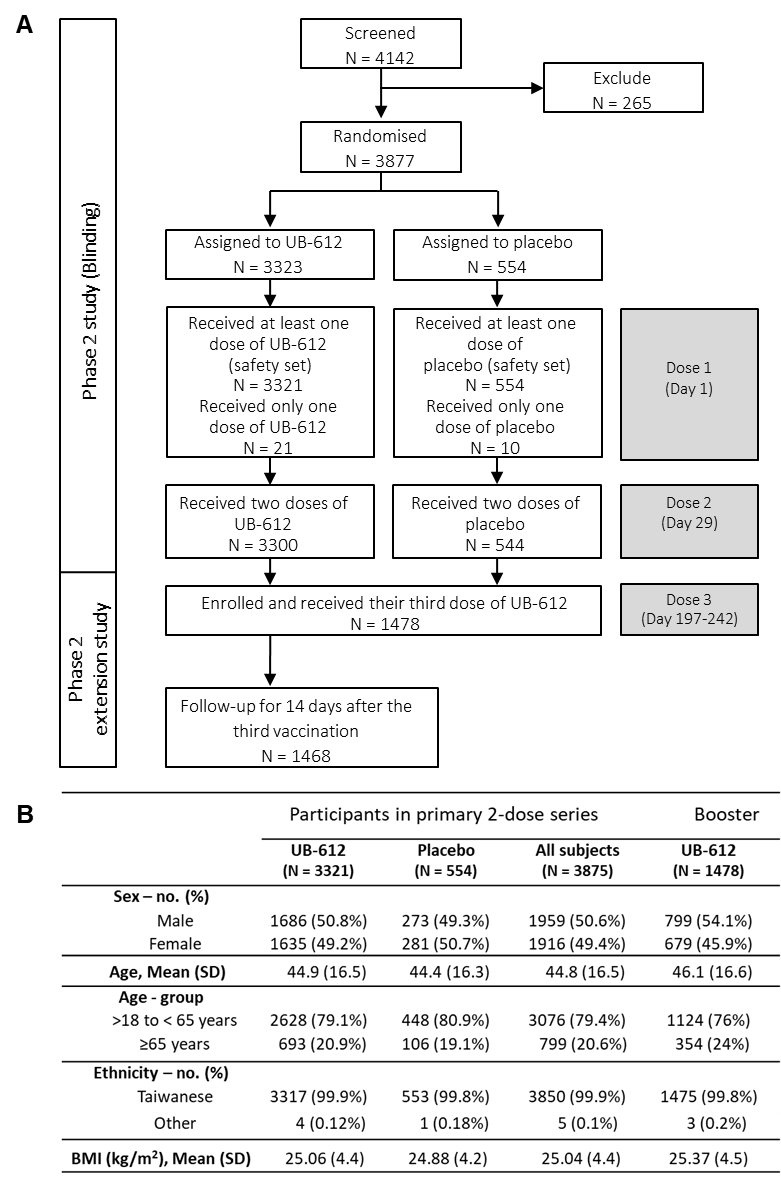

The study design of the phase II primary 2-dose series (100 μg dose; 28 days apart) of UB-612; and the extension study of booster vaccination [NCT04773067] conducted between Oct. 16, 2021 and Apr. 16, 2022. **(A)** Of the primary series (n = 3875), a total of 1,478 participants (aged at 18-85 years) were enrolled to receive the booster third-dose of 100 μg UB-612; **(B)** the characteristics of the study participants in the primary and booster series.

**Figure** **S2. Incidence of adverse effects in the phase II primary 2-dose and extended booster third-dose series**

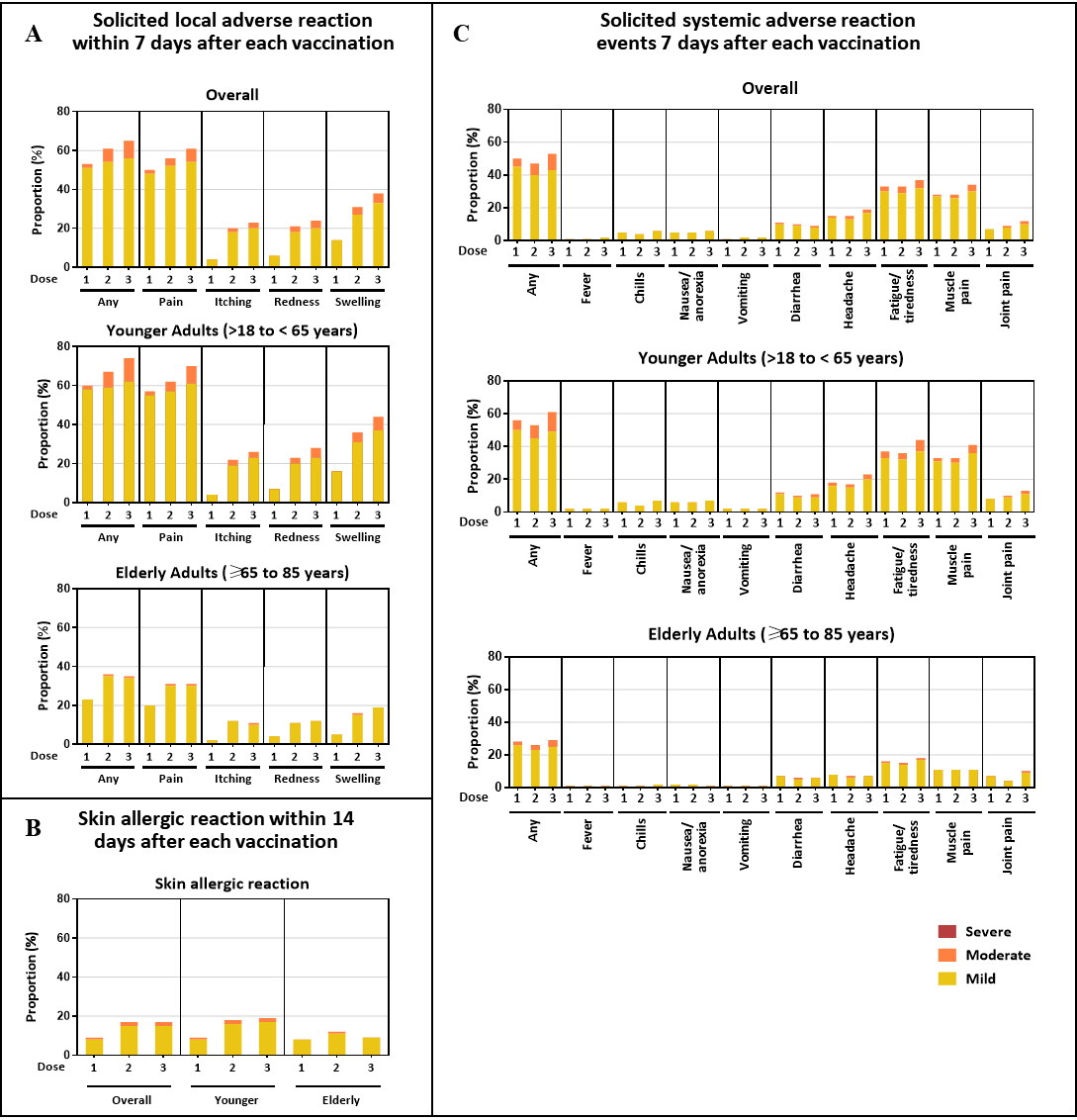

**(A)** Solicited local adverse reaction within 7 days after each vaccination. **(B)** Skin allergic reaction within 14 days after each vaccination. **(C)** Solicited systemic adverse reaction events 7 days after each vaccination (Doses 1 and 2 in the primary series; Dose 3 as a booster)

**Figure S3. Phase-2 immunogenicity overview (antigenic and functional) on**

**homologous boosting**

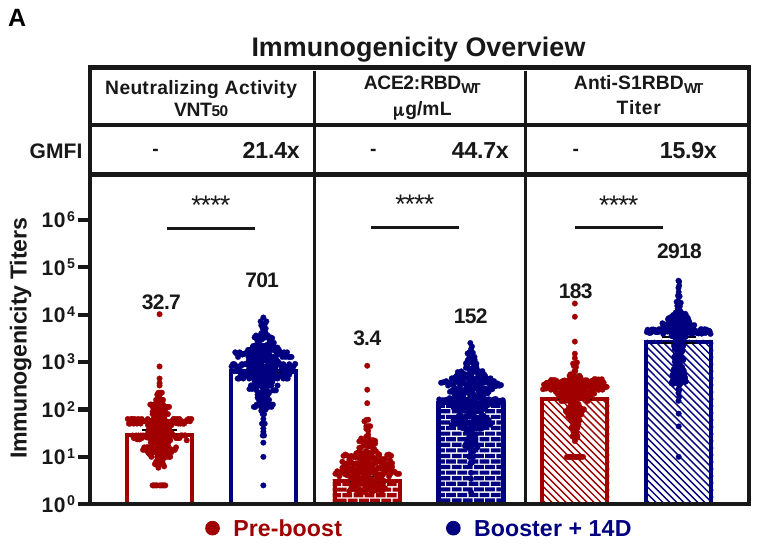

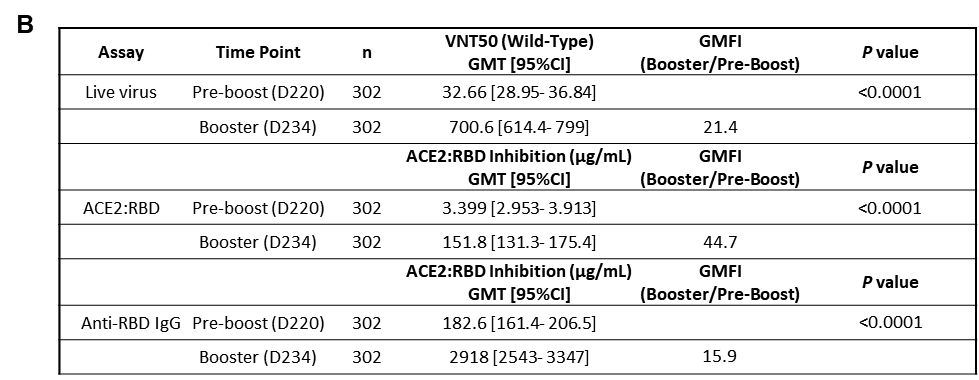

Immunogenicity overview are presented in **(A)** antigenic S1-RBD_WT_ binding, ACE2:RBDWT binding inhibition, and anti-WT viral-neutralizing activity VNT_50_ and **(B)** the **s**ummary of geometric mean titer (GMT) with 95% CI. A total of 302 participants (n = 208 for aged 18-65 years; n = 94 for aged 65-85 years) received a booster 3^rd^-dose. The serum samples of 302 participants were collected at the indicted time points, Days 197 to 242 (the pre-booster day) and Days 211 to 256 (14 days post-booster), and tested for neutralizing antibody levels that inhibit 50% of live SARS-CoV-2 wild-type, expressed as VNT_50_ (WT, Wuhan strain) (functional), the inhibitory titers against S1-RBD binding to ACE2 by ELISA, expressed as μg/mL (functional), and anti-S1-RBD IgG antibody titers by ELISA (antigenic). Statistical analysis was performed by the Student’s t-test (ns *p*>0.05, **** *p*<0.0001).

**Figure S4. Viral-neutralizing titers against live SARS-CoV-2 wild type (Wuhan) and Delta**

**variant (VNT_50_), and pseudo SARS-CoV-2 wild type (Wuhan) and Omicron BA.1 variant (pVNT_50_) after the booster third-dose in the Phase-1 trial***

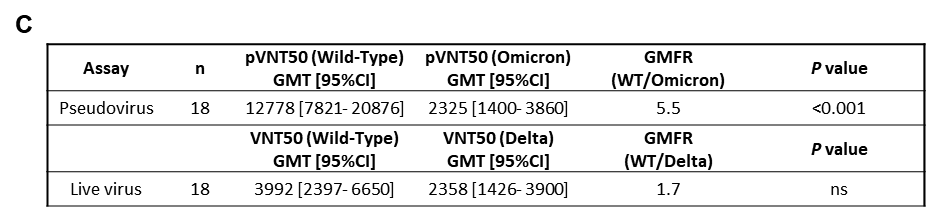

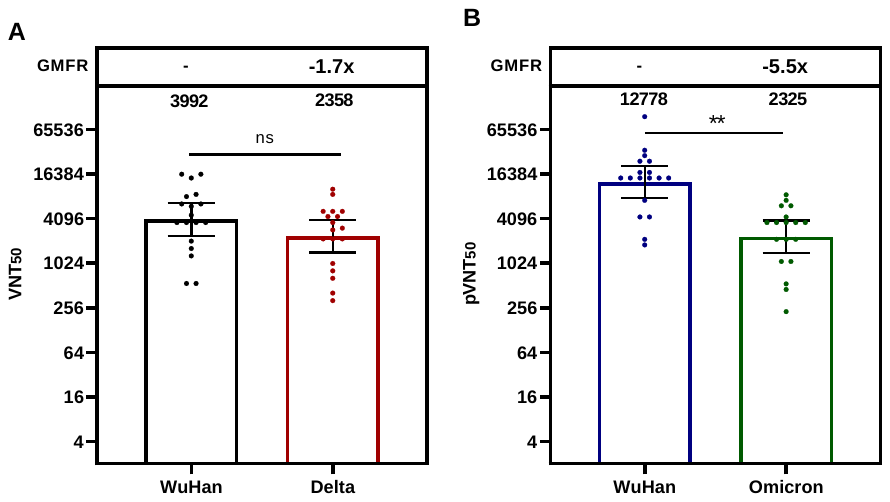

Geometric mean titers (GMT) at 50% viral-neutralization observed 14 days after the booster third-dose of 100-µg administered at mean Day 286 (Days 255-316) after the primary 2-dose series (Days 0 and 28) of the 196-day phase I trial. **(A)** In the participants of the 100-µg group (n = 18) with healthy adults aged at 20-55 years, the post-booster VNT_50_ titer reached at 3,992 against live SARS-CoV-2 Wuhan wild-type, and at 2,358 against live Delta variant. **(B)** Similarly, unusually high post-booster pVNT_50_ against Wuhan wild-type pseudovirus at 12,778, and at 2,325 against Omicron BA.1 variant. **(C)** Summary of geometric mean titer (GMT) with 95% CI are presented for plots shown in Figures **(A)** and **(B)**.

***** Adapted with permission from *J. Clin. Invest.* 2022;132(10):e157707. <https://doi.org/10.1172/JCI157707>

**Supplemental Tables**

**Table S1. Comparison of post-booster viral-neutralizing antibody titers against SARS-CoV-2 wild-type (WT) and Delta variant by vaccines from different platforms***

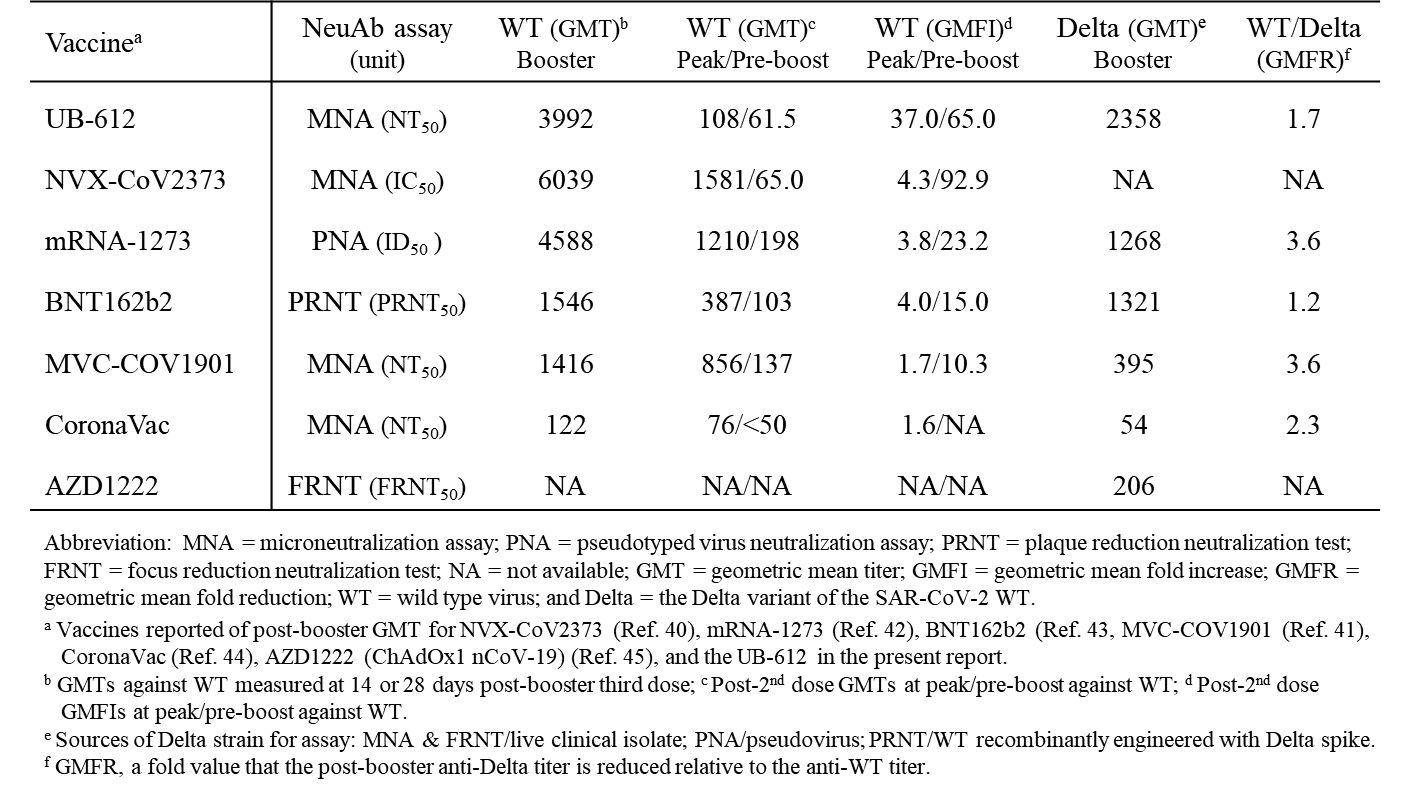

Abbreviation: MNA = Microneutralization assay; PNA = pseudotyped virus neutralization assay; PRNT = plaque reduction neutralization test; FRNT = focus reduction neutralization test; NA = not available; GMT = geometric mean titer; GMFI = geometric mean fold increase; GMFR = geometric mean fold reduction; WT = wild type virus; and Delta = the Delta variant of the SAR-CoV-2 WT.

^a^ Vaccine reported of post-booster GMT for NVX-CoV2373 (Ref. 40), mRNA-1273 (Ref. 42), BNT16b2 (Ref. 43), MVC-Cov1901 (Ref. 41), Corona Vac (Ref. 44), ADZ1222 (ChAdOx1 nCov-19) (Ref. 45), and UB-612 in the present report.

^b^ GMTs against WT measured at 14 or 28- days post-booster third dose.

^c^ Post-2^nd^ dose GMTs at peak/pre-booster against WT.

^d^ Post-2^nd^ dose GMFIs at peak/pre-booster against WT.

^e^ Sources of Delta strain for assay: MNA & FRNT/live clinical isolate: PNA/pseudovirus: PRNT/WT recombinantly engineered with Delta spike.

^f^ GMFR, a fold value that the post-booster anti-Delta titer is reduced relative to the anti-WT titer.

***** Reprinted with permission from *J. Clin. Invest.* 2022;132(10):e157707. <https://doi.org/10.1172/JCI157707>.

**Supplemental Methods**

**Vaccine product and placebo**. UB-612 vaccine used in the present phase II extension booster vaccination is a multitope vaccine designed to activate both humoral and cellular responses. For SARS-CoV-2 immunogens, UB-612 combines a CHO-expressed S1-RBD-sFc fusion protein (Wuhan strain) and a mixture of synthetic T helper (Th) and cytotoxic T lymphocyte (CTL) epitope peptides, which are selected from immunodominant M, S2 and N regions known to bind to human major histocompatibility complexes (MHC) I and II. The preparation of UB-612 vaccine product consists of compounding, filtration, mixing, and filling operations. Before addition of the subunit protein S1-RBD-sFc, the individual components of the vaccine are filtered through a 0.22 micron membrane filter, including the peptide solution (2 µg/mL), CpG1, a proprietary oligonucleotide (ODN), solution (2 µg/mL), 10X protein buffer containing 40 mM Histidine, 500 mM Arginine and 0.6% Tween 80, 20% sodium chloride stock solution. After sequentially addition of each component, the S1-RBD-sFc fusion protein and peptides are formulated with components described as above to form a protein-peptide complex and then is adsorbed to aluminum phosphate (Adju-Phos®) adjuvant. The last step would be addition of water for injection containing the 2- phenoxyethanol preservative solution to make final drug product at 200 µg/mL. The UB-612 vaccine product is stored at 2 to 8 ^o^C. Placebo used in the Phase-2 trial was sterile 0.9% normal saline.

**Viral-neutralizing antibody titers against SARS-CoV-2 wild-type and variants by CPE based live virus neutralization assay.** Neutralizing antibody titers were measured by CPE-based live virus neutralization assay using Vero-E6 cells challenged with wild type (SARS-CoV-2-Taiwan-CDC#4, Wuhan) and Delta variant (SARS-CoV-2-Taiwan-CDC#1144, B.1.617.2), which was conducted in a BSL-3 lab at Academia Sinica, Taiwan. Vero-E6 (ATCC® CRL-1586) cells were cultured in DMEM (Hyclone) supplemented with 10% fetal bovine serum (FBS, Gibco) and 1x Penicillin-Streptomycin solution (Thermo) in a humidified atmosphere with 5% CO_2_ at 37°C. The 96-well microtiter plates are seeded with 1.2×10^4^ cells/100 μL/well. Plates are incubated at 37^o^ C in a CO_2_ incubator overnight. The next day tested sera were heated at 56 °C for 30 min to inactivate complement, and then diluted in DMEM (supplemented with 2% FBS and 1x Penicillin/Streptomycin). Serial 2-fold dilutions of sera were carried out for the dilutions. Fifty μL of diluted sera were mixed with an equal volume of virus (100 TCID50) and incubated at 37°C for 1 hr. After removing the overnight culture medium, 100 μL of the sera-virus mixtures were inoculated onto a confluent monolayer of Vero-E6 cells in 96-well plates in triplicate. After incubation for 4 days at 37 °C with 5% CO_2_, the cells were fixed with 10% formaldehyde and stained with 0.5% crystal violet staining solution at room temperature for 20 min. Individual wells were scored for CPE as having a binary outcome of ‘infection” or ‘no infection’. Determination of SARS-CoV-2 virus specific neutralization titer was to measure the neutralizing antibody titer against SARS-CoV-2 virus based on the principle of VNT50 titer (≥50% reduction of virus-induced cytopathic effects). Virus neutralization titer of a serum was defined as the reciprocal of the highest serum dilution at which 50% reduction in cytopathic effects are observed and results are calculated by the method of Reed and Muench.

**Neutralizing titers against Omicron BA.1/BA.2/BA.5 by pesudovirus assay.** Neutralizing antibody titers were measured by neutralization assay using HEK-293T-ACE2 cells challenged with SARS-CoV-2 pseudovirus variants. The study was conducted in a BSL2 lab at RNAi core, Biomedical Translation Research Center (BioTReC), Academia Sinica. Human embryonic kidney (HEK-293T/17; ATCC® CRL-11268^TM^) cells were obtained from the American Type Culture Collection (ATCC). Cells were cultured in DMEM (Gibco) supplemented with 10% fetal bovine serum (Hyclone) and 100 U/mL of Penicillin-Streptomycin solution (Gibco), and then incubated in a humidified atmosphere with 5% CO2 at 37 °C. HEK-293T-ACE2 cells were generated by transduction of VSV-G pseudotyped lentivirus carrying human ACE2 gene. To produce SARS-CoV-2 pseudoviruses, a plasmid expressing C-terminal truncated wild-type Wuhan-Hu-1 strain SARS-CoV-2 spike protein (pcDNA3.1-nCoV-SΔ18) was co-transfected into HEK-293T/17 cells with packaging and reporter plasmids (pCMVΔ8.91, and pLAS2w.FLuc.Ppuro, respectively) (BioTReC, Academia Sinica), using TransIT-LT1 transfection reagent (Mirus Bio). Site-directed mutagenesis was used to generate the Omicron BA.1, BA.2, and BA.4/BA.5 variants by changing nucleotides from Wuhan-Hu-1 reference strain. For BA.1 variant, the mutations of spike protein are A67V, Δ69-70, T95I, G142D/Δ143-145, Δ211/L212I, ins214EPE, G339D, S371L, S373P, S375F, K417N, N440K, G446S, S477N, T478K, E484A, Q493R, G496S, Q498R, N501Y, Y505H, T547K, D614G, H655Y, N679K, P681H, N764K, D796Y, N856K, Q954H, N969K, L981F. For BA.2 variant, the mutations of spike protein are T19I, L24S, Δ25-27, G142D, V213G, G339D, S371F, S373P, S375F, T376A, D405N, R408S, K417N, N440K, S477N, T478K, E484A, Q493R, Q498R, N501Y, Y505H, D614G, H655Y, N679K, P681H, N764K, D796Y, Q954H, N969K. For BA.4/5 variant, the mutations of spike protein are T19I, L24S, Δ25-27, Δ69-70, G142D, V213G, G339D, S371F, S373P, S375F, T,376A, D405N, R408S, K417N, N440K, L452R, S477N, T478K, E484A, L486V, Q493, Q498R, N501Y, Y505H, D614G, H655Y, N679K, N764K, D796Y, N856K, and Q954H, & L969K.

Indicated plasmids were delivered into HEK-293T/17 cells by using TransITR-LT1 transfection reagent (Mirus Bio) to produce different SARS-CoV-2 pseudoviruses. At 72 hours post-transfection, cell debris were removed by centrifugation at 4,000 xg for 10 minutes, and supernatants were collected, filtered (0.45 μm, Pall Corporation) and frozen at −80 °C until use. HEK-293-hACE2 cells (1x10^4^ cells/well) were seeded in 96-well white isoplates and incubated for overnight. Tested sera were heated at 56°C for 30 min to inactivate complement, and diluted in medium (DMEM supplemented with 1% FBS and 100 U/ml Penicillin/Streptomycin), and then 2-fold serial dilutions were carried out for a total of 8 dilutions. The 25 μL diluted sera were mixed with an equal volume of pseudovirus (1,000 TU) and incubated at 37 °C for 1 hr before adding to the plates with cells. After 1-hr incubation, the 50 μL mixture added to the plate with cells containing with 50 μL of DMEM culture medium per well at the indicated dilution factors. On the following 16 hours incubation, the culture medium was replaced with 50 μL of fresh medium (DMEM supplemented with 10% FBS and 100 U/ml Penicillin/Streptomycin). Cells were lysed at 72 hours post-infection and relative light units (RLU) was measured by using Bright-GloTM Luciferase Assay System (Promega). The luciferase activity was detected by Tecan i-control (Infinite 500). The percentage of inhibition was calculated as the ratio of RLU reduction in the presence of diluted serum to the RLU value of virus only control and the calculation formula was shown below: (RLU ^Control^ - RLU ^Serum^) / RLU ^Control^. The 50% protective titer (NT50 titer) was determined by Reed and Muench method.

**Inhibition of RBDWT binding to ACE2 by ELISA**. The 96-well ELISA plates were coated with 2 µg/mL ACE2-ECD-Fc antigen (100 μL/well in coating buffer, 0.1M sodium carbonate, pH 9.6) and incubated overnight (16 to 18 hr) at 4 °C. Plates were washed 6 times with Wash Buffer (25-fold solution of phosphate buffered saline, pH 7.0-7.4 with 0.05% Tween 20, 250 μL/well/wash) using an Automatic Microplate Washer. Extra binding sites were blocked by 200 μL/well of blocking solution (5 N HCl, Sucrose, Triton X-100, Casein, and Trizma Base). Five-fold dilutions of immune serum or a positive control (diluted in a buffered salt solution containing carrier proteins and preservatives) were mixed with a 1:100 dilution of RBDWT-HRP conjugate (horseradish peroxidase-conjugated recombinant protein S1-RBD-His), incubated for 30±2 min at 25±2 °C, washed and TMB substrate (3,3’,5,5’-tetramethylbenzidine diluted in citrate buffer containing hydrogen peroxide) is added. Reaction is stopped by stop solution (diluted sulfuric acid, H_2_SO_4,_ solution, 1.0 M) and the absorbance of each well is read at 450nm within 10 min using the Microplate reader (VersaMax). Calibration standards for quantitation ranged from 0.16 to 2.5 μg/mL. Samples with titer value below 0.16 μg/mL were defined as being half of the detection limit. Samples with titer exceed 2.5 μg/mL were further diluted for reanalysis.

**Anti-S1-RBDWT binding IgG antibody by ELISA.** The 96-well ELISA plates were coated with 2 µg/mL recombinant S1-RBDWT-His protein antigen (100 µL/well in coating buffer, 0.1 M sodium carbonate, pH 9.6) and incubated overnight (16 to 18 hr) at room temperature. One hundred μL/well of serially diluted serum samples (diluted from 1:20, 1:1,000, 1:10,000 and 1:100,000, total 4 dilutions) in 2 replicates were added and plates are incubated at 37 °C for 1 hr. The plates were washed six times with 250 μL Wash Buffer (PBS-0.05% Tween 20, pH 7.4). Bound antibodies were detected with HRP-rProtein A/G at 37 ^o^C for 30 min, followed by six washes. Finally, 100 μL/well of TMB (3,3’,5,5’-tetramethylbenzidine) prepared in Substrate Working Solution (citrate buffer containing hydrogen peroxide) was added and incubated at 37 ^o^C for 15 min in the dark, and the reaction stopped by adding 100 μL/well of H_2_SO_4,_ 1.0 M. Sample color developed was measured on ELISA plate reader (Molecular Device, VersaMax). UBI® EIA Titer Calculation Program was used to calculate the relative titer. The anti-S1-RBD antibody level is expressed as Log_10_ of an end point dilution for a test sample (SoftMax Pro 6.5, Quadratic fitting curve, Cut-off value 0.248).

**T cell responses by ELISPOT.** Human peripheral blood mononuclear cells (PBMCs) were used in the detection of the T cell response. For the booster-series third-dose series extension study, ELISpot assays were performed using the human IFN-γ/IL-4 FluoroSpot^PLUS^ kit (MABTECH). Aliquots of 250,000 PBMCs were plated into each well and stimulated, respectively, with 10 μg/mL (each stimulator) of RBD-WT+Th/CTL, Th/CTL, or Th/CTL pool without UBITh1a (CoV2 peptides), and cultured in culture medium alone as negative controls for each plate for 24 hours at 37 °C with 5% CO2. The analysis was conducted according to the manufacturer’s instructions. Spot-forming units (SFU) per million cells was calculated by subtracting the negative control wells.

**Intracellular cytokine taining (ICS).** Intracellular cytokine staining and flow cytometry was used to evaluate CD4^+^ and CD8^+^ T cell responses. PBMCs were stimulated, respectively, with S1-RBD-His recombinant protein plus with Th/CTL peptide pool, Th/CTL peptide pool only, CoV2 peptides, PMA + Inonmycin (as positive controls), or cultured in culture medium alone as negative controls for 6 hours at 37°C with 5% CO_2_. Following stimulation, cells were washed and stained with viability dye for 20 minutes at room temperature, followed by surface stain for 20 minutes at room temperature, cell fixation and permeabilization with the BD cytofix/cytoperm kit (Catalog # 554714) for 20 minutes at room temperature, and then intracellular stain for 20 minutes at room temperature. Intracellular cytokine staining of IFN-γ, IL-2 and IL-4 was used to evaluate CD4^+^ T cell response. Intracellular cytokine staining of IFN-γ, IL-2, CD107a and Granzyme B was used to evaluate CD8^+^ T cell responses. Upon completion of staining, cells were analyzed in a FACSCanto II flow cytometry (BD Biosciences) using BD FACSDiva software.

**Statistics**. For the phase II extension booster vaccination study, the immunogenicity results for Geometric Mean Titer (GMT) are presented with the 95% confidence intervals. Statistical analyses were performed using SAS® Version 9.4 (SAS Institute, Cary, NC, USA) or Wilcoxon sign rank test. Spearman correlation was used to evaluate the monotonic relationship between non-normally distributed data sets. For the phase II primary 2-dose series, the sample size of the trial design meets the minimum safety requirement of 3000 study participants in the vaccine group with healthy adults, as recommended by the US FDA and WHO.

US Food and Drug Administration, Emergency use authorization for vaccines to prevent COVID-19: Guidance for industry, <https://downloads.regulations.gov/FDA-2020-D-1137-0019/attachment_1.pdf>;

WHO Guidelines on clinical evaluation of vaccines: regulatory expectations, <https://cdn.who.int/media/docs/default-source/prequal/vaccines/who-trs-1004-web-annex-9.pdf?sfvrsn=9c8f4704_2&download=true>.

**Supplemental Appendices** (available upon request)

Appendix 1

Phase II study V-205 protocol

Appendix 2

Phase II study V-205 IRB approval letter 1

Phase II study V-205 IRB approval letter 2

Phase II study V-205 IRB approval letter 3

Appendix 3

Phase II study V-205 Informed Consent Form (ICF)

Appendix 4

CONSORT checklist
